# An atlas of genetic effects on the monocyte methylome across European and African populations

**DOI:** 10.1101/2024.08.12.24311885

**Authors:** Wanheng Zhang, Chuan Qiu, Xiao Zhang, Zichen Zhang, Kuan-Jui Su, Zhe Luo, Minghui Liu, Bingxin Zhao, Lang Wu, Qing Tian, Hui Shen, Chong Wu, Hong-Wen Deng

## Abstract

**Background:** Genetic regulation of DNA methylation in immune cells may mediate complex disease risk. However, current epigenomic studies are constrained by microarray CpG coverage, mixed-cell tissues, and limited representation of diverse ancestries. Thus, we generated a whole-genome, multi-ancestry atlas of genetic effects on the purified monocyte methylome.

**Methods:** We first performed whole-genome bisulfite sequencing (WGBS) of purified peripheral blood monocytes and whole-genome sequencing (WGS) from 160 African American (AA) and 298 European American (EA) participants, profiling around 25 million CpG sites. Next, we identified *cis*-methylation quantitative trait loci (meQTLs), estimated cis heritability, and evaluated replication against large external meQTL resources. We further trained population-specific DNAm imputation models and applied them to methylome-wide association studies (MWAS) of 41 traits using genome-wide association study summary statistics from the Million Veteran Program. Type 2 diabetes signals were further evaluated using Mendelian randomization and Bayesian colocalization. We also conducted exploratory trans-meQTL mapping.

**Results:** We identified 1,480,064 and 1,527,480 CpG sites with at least one cis-meQTL in AA and EA populations, respectively, including 543,869 shared sites and extensive population-specific regulation attributable to both allele-frequency differences and effect-size heterogeneity. Cis-meQTL effects replicated robustly in external datasets: effect sizes correlated strongly with prior studies (EA Pearson’s r = 0.76; 90.8% concordant directions; AA Pearson’s r = 0.71; 86.6% concordant directions). We built DNAm prediction models with *cis*-h^2^ > 0.01 for 2,677,714 CpG sites in AA and 1,976,046 CpG sites in EA, achieving mean cross-validated prediction R^2^ of 0.20 and 0.18. Across 41 traits, MWAS 23,650 significant methylation-phenotype associations (2,116 in AA and 21,534 in EA), of which ∼98% were not interrogated by Illumina 450K/EPIC arrays. For type 2 diabetes, MWAS identified 20 CpG sites in AA and 4,023 CpG sites in EA, with substantial support from Mendelian randomization and colocalization. Exploratory trans-meQTL mapping detected widespread long-range associations, with limited cross-study overlap but high directional concordance among shared signals.

**Conclusions:** This whole-genome, monocyte-resolved, multi-ancestry methylome atlas and accompanying imputation resource expand interpretable methylation variation beyond array-based studies and enable multi-ancestry integration of genetic, epigenetic, and genome-wide association study data to prioritize immune-cell regulatory mechanisms for complex disease.

## Background

Unraveling the functional consequences of genetic variation in complex human diseases remains a central challenge in human genetics. While genome-wide association studies (GWAS) have identified numerous single nucleotide polymorphism (SNP)-trait associations across a wide range of complex diseases and traits,^1^ the molecular mechanisms through which these variations contribute to disease risk remain largely elusive.^2,3^ This knowledge gap hinders the identification of therapeutic targets^4^ and underscores the importance of exploring multi-omics biomarkers^5^. Among these, DNA methylation (DNAm), a relatively stable epigenetic modification, has emerged as a critical molecular biomarker in this endeavor. DNAm not only provides insights into the current state of human health^6^ but also facilitates the estimation of epigenetic age^7–9^ and partially captures the impact of environment and lifestyle exposures^10^ on disease pathogenesis.

Recognizing its importance, extensive efforts have been devoted to generating comprehensive, population-scale datasets of the human genome and methylome.^11–19^ However, these pursuits have been constrained by several key limitations. Firstly, most epigenomic studies rely on Illumina BeadChip platforms ^20^, which target only 450,000 or 935,000 predefined CpG sites— capturing only ∼1.5-3% of the ∼30 million CpG sites in the human genome ^21–24^. This limited coverage restricts our ability to fully explore the methylome’s extensive landscape. Secondly, the majority of DNAm studies are based on whole blood ^25–28^ or bulk tissue samples^29–31^, incorporating the estimated cell type proportions as covariates in subsequent analysis^32,33^. This approach inherently limits the examination of cell type-specific methylation patterns, which are crucial for elucidating disease mechanisms.^34,35^ Monocytes, in particular, play central roles in various physiological and pathological processes, including bone remodeling^36–38^, neurodegenerative disorders such as Alzheimer’s disease^39,40^, inflammatory conditions like rheumatoid arthritis^41^, cardiovascular diseases^42^, and cancer^43^, making monocytes an ideal model for investigating how genetic variation shapes DNAm patterns relevant to complex diseases. Thirdly, both DNAm and genetic variations can be ancestry-specific^44–46^, highlighting the critical need to study DNAm across diverse populations to comprehensively characterize these relationships.

To overcome these limitations, we performed whole genome bisulfite sequencing (WGBS) on purified monocyte DNA and whole genome sequencing (WGS) on blood-derived DNA from 160 African Americans (AA) and 298 European Americans (EA) participants in the Louisiana Osteoporosis Study (LOS)^47^. Our comprehensive, multi-ancestry approach encompassed several key analyses. For each ancestry group, we conducted methylation quantitative trait loci (meQTLs) analyses to identify genetic variants associated with DNAm in both *cis*- and *trans*-regions, and compared findings across populations to uncover shared and ancestry-specific regulatory architecture. Further, we estimated *cis*-heritability (*cis*-h^2^) of DNAm to quantify the proportion of methylation variance attributable to *cis*-acting genetic variation. To investigate the connection between DNAm and complex traits, we conducted methylome-wide association studies (MWAS) using a two-step instrumental variable regression framework^48,49^. First, we developed ancestry-specific DNAm imputation models using penalized regression. Second, we tested the association between the predicted DNAm and 41 phenotypes using GWAS summary statistics from the Million Veteran Program (MVP) dataset ^50^ for both EA and AA groups. We further analyzed Type 2 diabetes (T2D) results and validated them through colocalization and Mendelian randomization (MR) analyses. This multi-faceted approach provides novel insights into the genetic regulation of monocyte-specific DNAm and its relevance to complex traits across diverse ancestries. An overview of the study design is shown in Fig. 1.

**Fig. 1.**
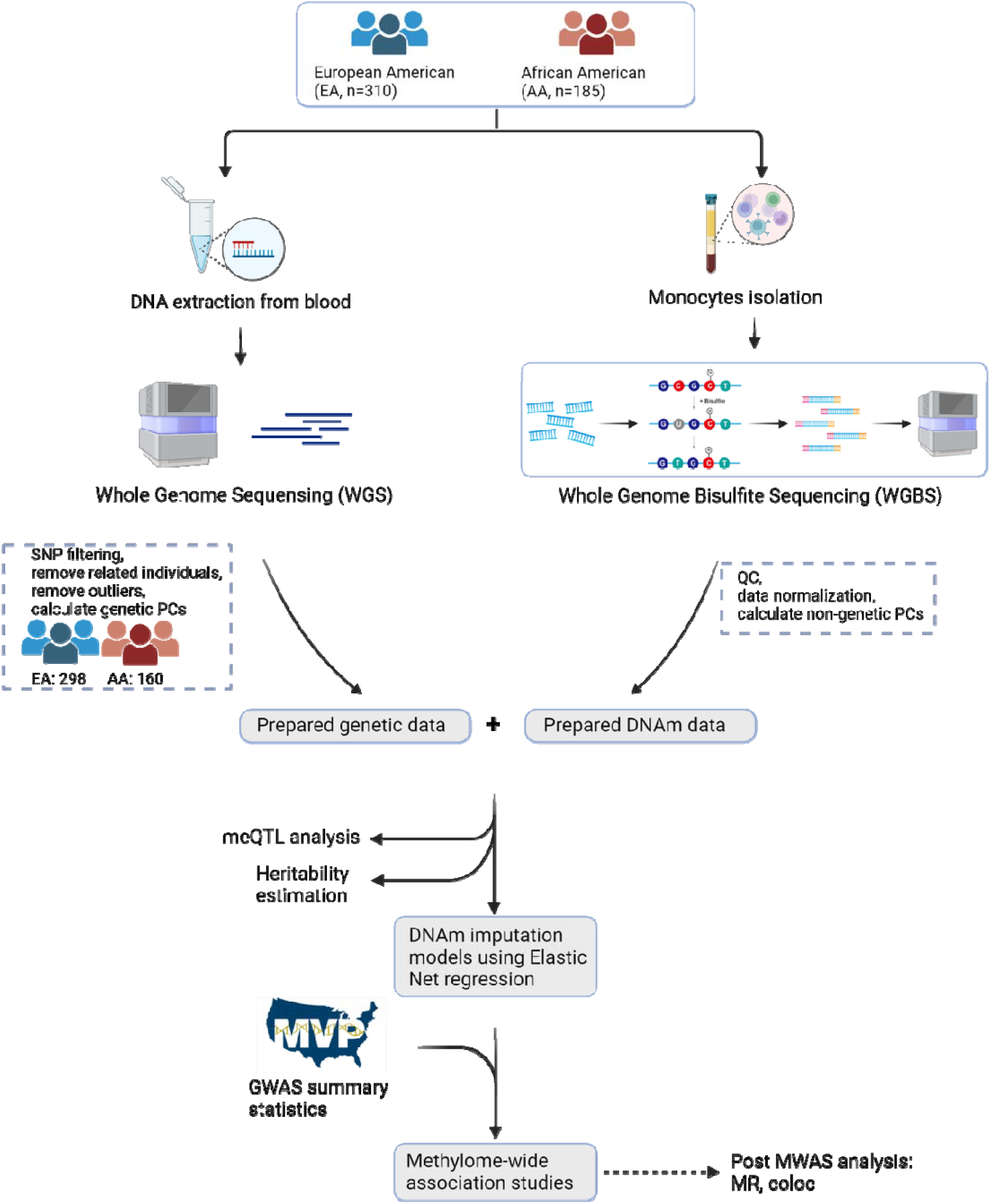
**Overview of the data collection, preparation, and analysis pipeline.**

## Methods

### Study subjects

We recruited a total of 495 male subjects (185 AA and 310 EA), aged 20–51 years (mean: 37.2 ± 8.8 years) from our ongoing Tulane LOS cohort. We excluded individuals with preexisting conditions affecting bone development and immune function, including: (1) cerebral vascular disease, (2) diabetes mellitus, except for easily controlled, noninsulin dependent cases, (3) chronic renal or liver failure, (4) chronic lung disease, (5) chronic obstructive pulmonary disease, (6) any metabolic or inherited bone diseases (e.g., hyper/hypoparathyroidism, Paget’s disease, osteomalacia, osteogenesis imperfecta, and hypochondrogenesis), (7) collagen disorder (e.g., rheumatoid arthritis, except for minor cases that involve only hand joint and wrist), (8) chronic gastrointestinal disease, (9) alcohol abuse, (10) treatment with corticosteroid or anticonvulsant therapy for more than 6 months duration, (11) antibiotic usage, (12) gastroenteritis, (13) major surgeries, (14) intercontinental travel in the past 3 months, (15) autoimmune or autoimmune-related diseases (e.g., multiple sclerosis), (16) immune-deficiency conditions (e.g., human Immunodeficiency Virus (HIV) infection), (17) haematopoietic and lymphoreticular malignancies (e.g., leukaemias), (18) active periods of asthma, or (19) influenza, infected within one week of recruitment. All recruited individuals signed an informed consent document prior to data and biosample collection. The study was approved by the Tulane University Institutional Review Board (IRB #: 10-184088).

### Whole Genome Sequencing (WGS)

Genomic DNA was extracted from 10 ml of fresh whole blood sample using the Gentra Puregene Blood Kit (Qiagen, USA). DNA concentration and quality were assessed with NanoDrop 2000 spectrophotometer, and samples were stored at -80 °C until further use. For each sample, 300 ng of genomic DNA was used to prepare sequencing libraries. Library preparation involved the generation of DNA Nanoballs (DNBs) through ligation-mediated polymerase chain reaction (LM-PCR), followed by single-strand separation, cyclization, and rolling circle amplification. WGS was performed on the DNBSEQ-500™ platform (BGI Americas, USA), with 150 bp paired-end reads in length. Sequencing reads were aligned to the GRCh38/hg38 human reference genome using Burrows-Wheeler Aligner (BWA, v0.7.12) ^51^. Genomic variants calling followed the Best Practices of the Genome Analysis Toolkit (GATK, v4.0.3) using HaplotypeCaller ^52,53^. The variant quality score recalibration (VQSR) was applied to obtain high-confidence variants ^52,53^.

### Isolation of monocytes and nucleic acids

In this study, we focused specifically on peripheral blood monocytes, which can act as osteoclast precursors and play important roles in regulating bone metabolism.^35^ Briefly, peripheral blood mononuclear cells (PBMCs) were firstly separated from ∼60 ml freshly collected peripheral blood, by a density gradient centrifugation method using Histopaque-1077 (Sigma-Aldrich, USA). The PBMCs were washed with 2 mM EDTA in PBS, and then resuspended in 0.5% bovine serum albumin (BSA) with 2 mM EDTA in PBS. Monocytes were subsequently isolated from the PBMCs with a Monocyte Isolation Kit II (Miltenyi Biotec, Germany) according to the manufacturer’s protocol. The kit uses negative selection to deplete unwanted cells (such as T and B cells) from PBMCs while preserving monocytes without surface-bound antibodies or beads. After negative selection, we validated monocyte purity using a multimodal workflow. First, for each sample, we prepared Wright–Giemsa–stained cytospins from the monocyte suspension and performed differential counts at 40–100× using standard cytomorphologic criteria (cell size, nuclear shape/chromatin, cytoplasmic basophilia/granularity) to distinguish monocytes from lymphocytes and granulocytes and to obtain an immediate purity estimate. Second, in three randomly selected samples, we performed single-cell RNA-seq, the cell-type clustering results demonstrated a predominant monocyte population based on CD14 transcripts (with concordant CD45 transcripts), yielding an estimated monocyte proportion of ∼85% (Additional file 1: Fig. S1), which is consistent with our prior flow-cytometry analysis^54,55^. Third, we estimated cell-type proportion in each sample using EPIC v1.1.7^56^, the deconvolution results indicated > 85% monocytes across all samples (Additional file 2: Table S1), corroborating the cytology and scRNA-seq-based purity assessments. The genomic DNA was extracted from the freshly isolated monocytes using the AllPrep DNA/RNA/miRNA Universal Kit (Qiagen, USA), following the manufacturer’s protocol. Samples were stored at -80 °C until further use.

### Whole Genome Bisulfite Sequencing (WGBS)

DNAm profiles were determined by WGBS according to previously published protocols.^57^ Briefly, 100 ng genomic DNA isolated from monocytes was fragmented to ∼250 bp by sonication using a Bioruptor (Diagenode, Belgium), followed by the blunt-ending, dA addition to 3’-end, and adaptor ligation. The ligated libraries were bisulfite converted using the EZ DNA Methylation-Gold kit (Zymo Research, USA) and sequenced on the Illumina HiSeq 4000 platform. Raw reads were filtered to remove adaptors, contaminants, and low-quality sequences. Reads with >10% unknown bases or >10% of bases with quality scores <20 were further excluded to obtain high-quality data. The cleaned reads were aligned to the GRCh37/hg19 human reference genome using BSMAP, then converted to GRCh38/hg38 using the UCSC LiftOver tool (https://genome.ucsc.edu/cgi-bin/hgLiftOver). The methylation level of each CpG site was determined by the ratio of the number of methylated reads to the total number of reads covering a particular cytosine site.

### Data preparation

We followed the GoDMC^11^ pipeline for the genotype data processing. Quality control (QC) was conducted on genotype data for chromosomes 1-22. SNPs that failed the Hardy-Weinberg equilibrium (*P*< 1 × 10^−6^) and had a minor allele frequency (MAF) < 0.05 were excluded. Duplicated SNPs and those with mismatched alleles were also removed, and indel alleles were recoded as “I” (insertion) and “D” (deletion). Next, we calculated the first 10 genetic principal components (PCs) using SNPs from HapMap3 reference panel, excluding those in regions of long-range linkage disequilibrium (LD) and retaining only SNPs with MAF > 0.2. Ancestry outliers, defined as individuals deviating more than 7 standard deviations from the mean along any PC, were excluded. Genetic PCs were then recalculated for downstream analyses. After QC, 7,803,505 SNPs in the AA cohort and 5,792,887 SNPs in the EA cohort remained for downstream analyses.

#### DNAm data normalization

To transform our DNAm data into a distribution that more closely approximates a Gaussian distribution, we applied a rank-based inverse normal transformation (INT)^60^. This method comprised two steps. First, the observations are transformed onto the probability scale using the empirical cumulative distribution function (ECDF). Second, the observations are transformed onto the real line, as Z-scores, using the probit function. After transformation, we removed CpG sites overlapping a known SNP within the CpG dinucleotide to avoid technical artifacts caused by allelic interference with methylation measurement (Additional file 1: Note. S1).

#### Co-variates

To ensure robust analysis and account for potential confounding factors, we implemented a comprehensive covariate adjustment strategy. First, we adjusted for key demographic and lifestyle factors, including age, body mass index (BMI), smoking status, and alcohol consumption. Second, to control for population structure and technical variation, we incorporated ten genetic PCs and ten non-genetic PCs. Genetic PCs were derived to capture the major axes of genetic variation. Non-genetic PCs were obtained by performing principal component analysis (PCA) on DNAm data, using 20,000 most variable CpG sites to extract the top ten components^11^. Incorporating non-genetic PCs derived from DNAm data has been shown to significantly improve the power to detect true associations^11^, while posing minimal risk of introducing collider bias.

### meQTL analyses

We performed a comprehensive analysis of both *cis*- and *trans*-meQTL mappings using the MatrixEQTL^61^ (R package), analyzing over 25 million normalized DNAm sites in AA (n = 160) and EA (n = 298) ancestries, separately. *Cis*-regions were defined as ±1 Mb from each CpG sites, with *cis*-acting meQTL (*cis*-meQTL) referring to SNPs located within this window and *trans*-acting meQTLs (*trans*-meQTLs) referring to SNPs located outside of this window. Using MatrixEQTL^61^, we identified *cis*-meQTL by testing the association between DNAm levels and genotype via linear regression, adjusting for potential confounders including age, BMI, smoking, alcohol consumption. Furthermore, we controlled for ten genetic and ten non-genetic principal components. Then, for each CpG sites *j*, the residual value, *y*_*ji*_, was regressed against each SNP *k*:

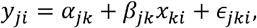

where genotype values *x*_*ki*_ were standardized to have a mean of zero and a standard deviation of one, *α*_*jk*_ was the intercept term, and *β*_*jk*_ was the effect estimate of each SNP *k* on each residualized CpG site *j*. We applied the Benjamini–Hochberg procedure to control the false discovery rate (FDR) at 0.01 for both *cis*- and *trans*-meQTLs.

### Simulation to evaluate the MAF–effect size relationship

Simulations were carried out using the genotype data in EA from the main study (n = 298). We drew a random set of *S* CpG sites and generated one phenotype for each site under a model where true SNP effects do not depend on MAF. For a CpG *s*, the cis window was defined as ±1 Mb around the CpG, and a single variant *c* (*s*) was chosen uniformly at random from the eligible SNPs in that window. Let *g*_*ik*_ ∈ {0,1,21 denote the additive genotype dosage for individual *i* at SNP *k*. let *p*_*k*_ denotes the MAF of *k*, and its sample variance is *v*_*k*_ ≈2*p*_*k*_ (1 - *p*_*k*_). Phenotypes followed

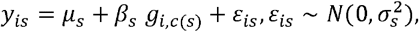

with effects drawn from a two-component mixture that is independent of MAF:

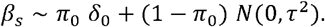

Unless stated otherwise we used *π*_0_ = 0.90 (10% non-zero effects) and *τ* = 0.40. To keep all CpGs on a comparable scale, we set 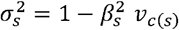 when *β*_*s*_ ≠ 0 and 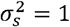 when *β*_s_ = 0; extremely rare draws yielding 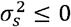were redrawn. Under this construction the site-specific *cis*-h^2^ for causal CpG (*β*_*s*_ ≠ 0 for their chosen *cis*-SNP) is

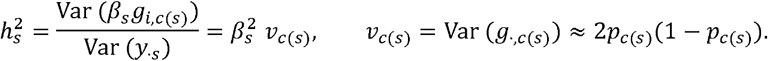

Each simulated CpG was tested against its selected SNP using a linear model in MatrixEQTL; false discovery rate was controlled at 0.05 by Benjamini–Hochberg.

### Replication studies

To validate our findings and assess the consistency of identified meQTLs across different populations and platforms, we conducted comprehensive replication analyses for both the AA and EA cohorts.

For the AA population, we compared our *cis*-meQTL results with those from the Genetic Epidemiology Network of Arteriopathy (GENOA)^18^ study. The GENOA study performed *cis*-meQTL mapping analysis on 961 African Americans, measuring DNAm using the Illumina HumanMethylation450 BeadChip platform. For replication analysis, we identified the subset of our *cis*-meQTLs that corresponded to CpG sites also measured in the GENOA study. We then compared the effect sizes and directions between the two studies for these overlapping sites. A successful replication was defined as a *cis*-meQTL association in the GENOA study with FDR < 0.05 and consistent direction of effect. We calculated Pearson’s correlation coefficient to quantify the concordance of effect sizes between studies.

For the EA population, we evaluated replication using data from the Genetics of DNA Methylation Consortium (GoDMC)^11^. The GoDMC study measured DNAm in whole blood using the Illumina HumanMethylation450 BeadChip across 27,750 European participants. This large-scale meta-analysis identified 420,509 methylation sites associated with genetic variants. Similar to our AA replication approach, we identified the subset of our EA *cis*-meQTLs that corresponded to CpG sites measured in the GoDMC study. We then compared effect sizes and directions between studies for these overlapping associations. A successful replication was defined as a *cis*-meQTL association in the GoDMC study with FDR < 0.05 and consistent direction of effect. We calculated Pearson’s correlation coefficient to assess concordance of effect size estimates between studies.

For *trans*-meQTL replication, we were limited to analyzing only the EA population since detailed *trans*-meQTL results were publicly available only from the GoDMC study, which exclusively contains European ancestry samples. Using the GoDMC data, we identified overlapping *trans*-meQTL associations with our EA cohort. We compared effect sizes using Pearson’s correlation and assessed the proportion of associations that maintained consistent direction of effect.

### Fine-mapping

To identify potentially causal variants influencing DNAm, we performed fine-mapping on *cis*-SNPs associated with each CpG site. This analysis specifically focused on CpG sites linked to at least one *cis*-meQTL in AA and EA populations, respectively. We used the *SuSiE* algorithm ^62^ from the *susieR* package in R to infer single effect components or credible sets for each CpG site and its associated variants. Each credible set was defined to have a 95% probability of encompassing at least one variant with a nonzero causal effect. We limited the number of credible sets per CpG site to a maximum of ten (L = 10), reflecting the assumption that up to ten causal variants may potentially regulate a single CpG site. To validate and compare fine-mapping results, we aggregated all credible sets identified by *SuSiE* for each CpG site by taking their union—thus capturing all variants with potential causal effects across the multiple credible sets (up to L=10). We then compared the sizes of these unified credible set between AA and EA populations to assess differences in fine-mapping resolution.

### *Cis*-h^2^

We employed Genome-wide Complex Trait Analysis (GCTA v1.94.1) ^63^ to estimate the h^2^ – the proportion of phenotypic variance attributable to genetic factors – for each CpG of ∼25 million CpG sites analyzed. Again, *cis*-regions were defined as within ±1 Mb of the CpG site, and heritability was estimated using only SNPs within these regions. GCTA constructs a genetic relationship matrix (GRM) from SNP data, which encapsulates the degree of genetic similarity between pairs of individuals. We then utilized this GRM within a restricted maximum likelihood (REML) framework to estimate h^2^. This approach allows for the separation of the phenotypic variance into components attributed to genetic variance (captured by the SNPs in the *cis*-regions) and residual variance. By focusing exclusively on *cis*-regional SNPs, our analysis specifically evaluated the local genetic contribution to DNAm variance at each CpG site, providing insights into the genetic architecture of epigenetic modifications.

### Comparison with Array-Based Platforms

To evaluate the added value of WGBS relative to array-based assays, we compared CpG coverage and downstream analytical performance using two commonly used platforms: the Infinium MethylationEPIC v2.0^64^, which contains over 935,000 CpG sites (900K), and the Infinium HumanMethylation450 BeadChip^65^, encompassing around 450,000 CpG sites (450K). Platform annotations were obtained from the InfiniumAnnotation resource at https://zwdzwd.github.io/InfiniumAnnotation. For each platform, we intersected the CpGs represented on the array with the CpGs profiled in our WGBS dataset and performed identical analyses on both sets.

### DNAm imputation model

In this study, we focused on CpG sites with estimated *cis*-h^2^ greater than 0.01. To enhance computational efficiency during model training, we adopted a 500-kb window around each target CpG site. For genotype data, following established practices ^66,67^, we excluded SNPs with a MAF < 1%, ambiguous strand orientation, insertions or deletions (indels), and SNPs absent in the LD reference panel derived from the 1000 Genomes Project ^68^. Our DNAm imputation model was constructed using a penalized regression approach, integrating methylation and genotype data within the defined *cis*-regions. Let *Y* denote the n-dimensional vector of methylation data for a given CpG site, and let ***X*** be the *n* ×*k* matrix of genotype dosages for *k cis*-SNPs associated with this CpG site:

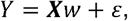

where is a *k* ×1 vector of SNP effect size to be estimated, and *ε* is the random error term with a mean of zero. To estimate *w*, we minimized the following penalized objective function *f* (*w*):

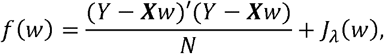

where *J_λ_*(*w*) represents a penalty term that regularizes the coefficients to prevent overfitting. Here, we used Elastic-net^69^ penalty, which combined both L1 and L2 regularization terms:

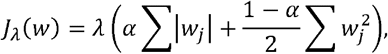

where *λ* is a tuning parameter that controls the overall strength of the penalty, *α* is the elastic-net mixing parameter, which is 0.5 in this study, that determines the trade-off between L1 penalty (lasso) and L2 penalty (ridge). *λ* is chosen via cross-validation.

The performance of the imputation models was evaluated by predictive *R*^2^, defined as the squared Pearson correlation coefficient between genetically predicted and directly measured DNAm data in nested cross-validation^70^. We only considered models with *R*^2^ > 0.01 in the subsequent analysis^48^.

### MWAS association testing in MVP database

In this section, we tested the association between genetically predicted DNAm levels and trait of interest. For phenotypes with only summary-level GWAS data available, we adopted the methodology described in previous studies^71,72^ to mitigate potential biases arising from discrepancies in LD matrices between reference panels and GWAS data. In our MWAS test, we estimated the effect size 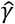 and its variance for the association between predicted DNAm and the phenotypes by:

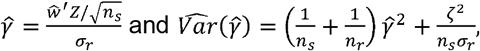

where *Z* is the vectors of z-scores from GWAS, 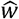 represents the estimated weights from the DNAm imputation model, *n*_*s*_ is the sample size of the GWAS data, *n*_*r*_ is the sample size of the population reference panel, 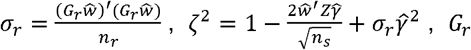, is the standardized genotype matrix of the population reference panel.

We applied this MWAS framework to 41 phenotypes available in the MVP^50^ database, for which GWAS summary statistics were available for both AA and EA ancestries. Details of these GWAS summary statistics are provided in Additional file 2: Table S2. Prior to association testing, we implemented QC on the GWAS summary statistics, including removal of duplicate entries, correction of strand orientation, and exclusion of ambiguous alleles, ensuring the integrity and reliability of our association analysis. To ensure robust methylation prediction, we excluded CpG sites with the number of non-zero weights from DNAm imputation models smaller than 10.

### Post-MWAS analyses

We compared our MWAS results with MR analysis in line with the methodology outlined by Zhao, et al.^73^. We selected a set of *cis*-meQTLs as the MR instrumental variables. We first restricted our analysis to the common set of variants that were shared by *cis*-meQTLs and the GWAS summary statistics. To avoid the potential issue of collinearity, we applied LD clumping and excluded *cis*-meQTLs with high correlation to index genetic variants (r^2^ > 0.001). Then, we removed variants that were not significant *cis*-meQTLs. To further ensure the validity of the instruments, we applied the Steiger filtering method ^74^ to exclude instrumental variables with potential reverse causality. Depending on the number of valid instrumental variables, we then applied either the Wald ratio method (for a single instrument) or inverse variance weighting^75^ (for multiple instruments) to assess the causal relationship between DNAm and the phenotype.

We also conducted Bayesian colocalization analyses^76^ on the significant CpG sites identified in our MWAS and MR analysis to estimate the posterior probability that the DNAm site and phenotype shared the same causal variant, using summary-level *cis*-meQTLs and GWAS data. Specifically, we used the coloc R package (v5.2.2), with its default setups in the coloc function, to estimate the posterior probability of both protein and phenotype being influenced by the same causal variant (i.e., the PPH4). We chose PPH4 > 0.8 as the threshold. DNAm satisfying this threshold would suggest a shared causal variant for the *cis*-meQTLs and GWAS associations.

## Results

### Data collection and processing

We recruited 495 male subjects aged 20-51 years (185 AA and 310 EA) from the ongoing Tulane LOS cohort^47^ with strict inclusion/exclusion criteria. Following standard QC for both genetic and DNAm data, 160 AA and 298 EA remained for analysis. The characteristics of the final cohort are summarized in Additional file 2: Table S3.

### Identification of *cis*-meQTL reveals methylation landscapes across populations through WGBS

Leveraging the comprehensive coverage of WGBS, we conducted an extensive analysis of *cis*-meQTLs across 24,750,692 CpG sites in AA (n=160) and 25,113,956 CpG sites in EA (n=298). We first computed meQTLs between each of these CpG sites and common SNPs within *cis*-regions, which identified 50,601,421 (*P* < 4.2 × 10^-6^) and 118,944,249 (*P* < 5.0 × 10^−5^) *cis*-meQTL associations in the AA and EA populations, respectively, when controlling FDR < 0.01. In total, 1,480,064 CpG sites in the AA and 1,527,480 in the EA population exhibited at least one significant *cis*-meQTL, corresponding to 6.0% and 6.1% of all tested sites, respectively. Among these, 543,869 CpG sites (36.7% of AA and 35.6% of EA) were shared between populations, indicating both common and population-specific genetic regulation of DNAm.

Our analysis uncovered several key patterns in the genetic architecture of methylation regulation. First, we observed a negative correlation between MAF and effect size in both AA (*r* = -0.33, *P* < 2.2 × 10^−16^) and EA (*r* = -0.34, *P* < 2.2 × 10^−16^) population (Fig. 2a), a pattern largely reflecting reduced statistical power to detect associations with low-frequency variants^77^. We further confirmed this through a simulation assuming MAF-independent effect sizes (see Methods; Additional file 1: Fig. S2). We also observed larger median effect sizes in AA (0.70) than in EA (0.46, *P* < 2.2 × 10^−16^ by Wilcoxon rank-sum test). These findings extend the principles observed in GWAS studies^78,79^ to methylation regulation, while highlighting important population differences. Second, we found that genetic variants closer to CpG sites typically showed stronger associations, with effect sizes decreasing as distance increased (Fig. 2b). This spatial relationship was confirmed using a non-parametric permutation test (AA: *ρ* = − 0.12; EA: *ρ* = − 0.13; permutation *P* < 0.0001). The relationship was consistent across both populations (Additional file 1: Fig. S3). Third, investigation of genomic context using Annotatr^80^ revealed that majority of CpG sites with *cis-*meQTLs were located in open sea regions (AA: 82.3%, EA: 81.8%), with smaller proportions in CpG islands (AA: 8.1%, EA: 7.7%) and shores/shelves (AA: 9.6%, EA: 10.4%). By comparison, among all tested CpG sites, 82.9% were located in open sea regions, 9.4% in shores/shelves, and 7.7% in islands. The enrichment of meQTLs in open sea regions aligns with previous findings^11^, suggesting that open sea CpGs are more susceptible to genetic regulation (Fig. 2c). Our findings are robust to MAF and significance cutoffs. Sensitivity analyses using multiple MAF thresholds (≥1% and ≥5%) and both FDR and fixed P-value cutoff at 1× 10^−6^ showed that all major patterns (MAF–effect-size relationship, distance decay, and genomic enrichment) remained unchanged (Additional file 1: Fig. S4 and Additional file 1: Fig. S5; Additional file 2: Table S4). Among the SNPs in all tested SNP-CpG pairs, the proportion of SNPs detected as significant *cis*-meQTLs dropped from 99.6% for common variants (MAF > 0.2) to 80.8% for low-frequency variants (0.01 < MAF ≤ 0.05) in EA, with a similar drop (95.8% to 75.1%) observed in AA (Additional file 1: Fig. S6).

**Fig. 2.**
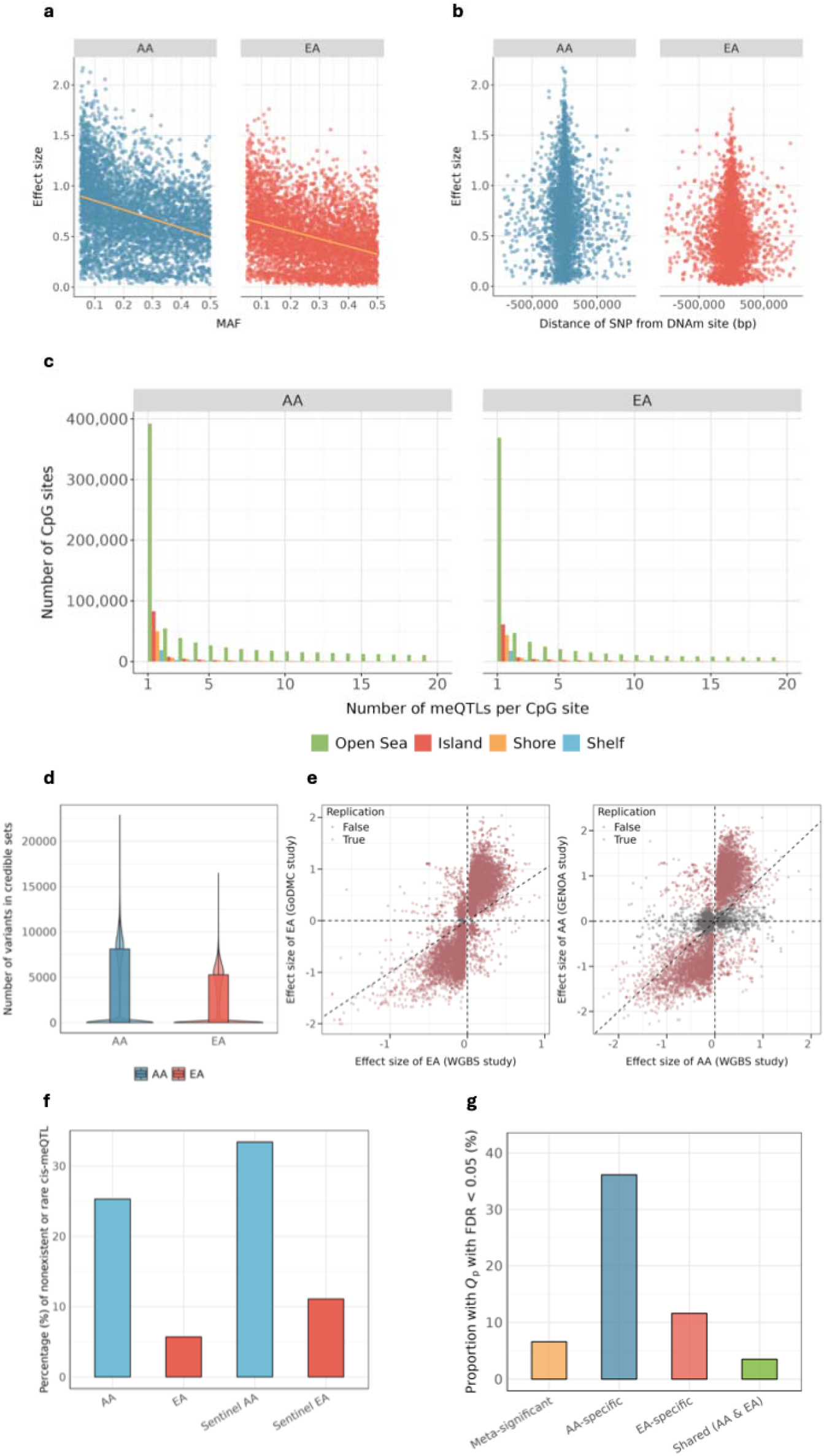
*cis*-meQTL and fine-mapping analysis across AA and EA populations. **(a)** Relationship between *cis*-meQTL effect sizes and MAF with regression trends shown in orange. **(b)** Effect size distribution as a function of distance between SNP and DNAm site in AA (left) and EA (right) populations, measured in base pairs. **(c)** Genomic distribution of CpG sites with *cis*-meQTLs. The bar chart categorizes CpG sites based on the number of *cis*-meQTLs identified, with annotations for four genomic regions: CpG islands, which are regions with a high CpG density; CpG shores, located within 2 kb of islands; CpG shelves, extending an additional 2 kb from shores; and Open Sea, which represents areas more distal to the islands, shores, and shelves. Each region is color-coded. **(d)** Fine-mapping results showing the distribution of credible set sizes from SuSiE analysis for CpG sites with significant *cis*-meQTLs in both populations. Box plots show median, quartiles, and 1.5×IQR whiskers; violin width indicates data density. **(e)** Replication effect sizes between WGBS study (EA, n = 298) and GoDMC (EA, n = 27,750). Effect sizes between WGBS study (AA, n = 160) and GENOA (AA, n = 961). **(f)** Percentage of nonexistent or rare *cis*-meQTLs across ancestries. It displays the proportion of identified *cis*-meQTLs that are absent or rare within the contrasting population from the 1000 Genomes Project. The four bars represent *cis*-meQTL associations for AA, EA, sentinel *cis*-meQTLs for AA and EA, respectively. Sentinel *cis*-meQTL is defined as the most significant SNPs for each CpG site. **(g)** The percentage of *cis*-meQTLs with significant effect-size heterogeneity, grouped by population specificity based on Cochran’s Q test (FDR < 0.05). Meta-significant: significant in meta-analysis; AA-specific or EA-specific: significant only in one ancestry; Shared: significant in both ancestries.

We then performed a down-sampling analysis to assess the impact of differential sample sizes on these discovery metrics. We randomly down-sampled the EA cohort to match the AA sample size (N = 160) and repeated the *cis*-meQTL mapping. This analysis yielded 47,986,816 *cis*-meQTLs in the down-sampled EA cohort (FDR < 0.01, *P* < 4.8 × 10^−6^), a substantial reduction from those identified in the full cohort, and this number is fewer than the associations detected in the AA cohort. This reduction was consistent at the CpG level, where the number of CpG sites with *cis*-meQTLs decreased to 908,106, and the number of shared CpG sites between populations dropped to 447,806. It confirms that the larger sample size was the primary driver of the higher *cis*-meQTL counts in full EA analysis. Importantly, all key characteristics of the *cis*-meQTLs, including the negative correlation between MAF and effect size (*r* = − 0.36 for AA and *r*= − 0.34 for EA), larger median effect size in AA (0.70 in AA and 0.64 in EA, *P* < 2.2 × 10^−16^ by the Wilcoxon rank-sum test), the negative correlation between effect sizes and genomic distances, and the decay of effect size with genomic distance, remained consistent in the down-sampled cohort (Additional file 1: Fig. S7).

To further leverage our multi-ancestry cohort and boost statistical power, we performed both fixed- and random-effects meta-analyses across the two populations for all common SNP-CpG pairs. The fixed-effects meta-analysis identified 129,811,518 significant *cis*-meQTLs (FDR < 0.01, *P*< 1.7 × 10^−5^). Among these, 25.0% were also significant in AA, 72.9% were significant in EA, and 78.1% were significant in at least one ancestry. Fixed-effects meta-analysis recovered the majority of population-specific findings, retaining 89.0% of the AA cohort, 88.8% of the EA cohort *cis*-meQTLs, and 86.4% in either population. Importantly, 21.9% of the associations identified by the meta-analysis were novel, having not been detected in either population-specific analysis alone, underscoring the gain in statistical power achieved by combining the cohorts (Additional file 1: Fig. S8a). The random-effects meta-analysis, which accounts for between-ancestry heterogeneity and penalizes associations showing high heterogeneity, recovered fewer *cis*-meQTLs (64.2% of AA and 48.7% of EA), and identified 23.5% novel associations (Additional file 1: Fig. S8b). These findings indicate a substantial proportion of *cis*-meQTLs exhibit genuine ancestry-specific effect.

To narrow down 95% credible sets of causal variants underlying significant *cis*-meQTL, we conducted population-specific fine-mapping for CpG sites with at least one *cis*-meQTL in AA and EA populations using SuSiE^62^. We found that the number of variants within the 95% credible sets was notably smaller for the AA population, with a median of 14 and an interquartile range (IQR) of 8,147, as opposed to the EA population, which exhibited a median of 28 and an IQR of 5,286 (Fig. 2d). This pattern remained consistent when the EA cohort was down-sampled to match the AA sample size, with a median of 29 (IQR = 5,489), implying this observation most likely reflects lower LD in AA ancestry populations (Additional file 1: Fig. S7d).

Our identified *cis*-meQTLs demonstrated robust replication when compared with previous studies (Fig. 2e). In the AA cohort, we compared our results with the GENOA AA cohort (N = 961). Due to the substantial difference in methylation site coverage between our WGBS approach (over 25 million CpG sites) and the 450K array platform used in GENOA, replication data were available for only 1,824,388 of our identified *cis*-meQTLs. We observed a strong correlation of effect sizes (Pearson’s r = 0.71, *P* < 2.2 × 10^−16^), with 86.6% of associations showing consistent direction of effect. At FDR < 0.05 (P cutoff = 3.5 × 10^−6^), 77.3% (1,410,251) of our discovery meQTLs in AA successfully replicated in the GENOA AA cohort. For the EA cohort, we compared our findings with the GoDMC study, which measured DNAm in whole blood using the Illumina HumanMethylation 450K BeadChip. Replication data were available for 534,938 of our identified *cis*-meQTLs in EA. Analysis revealed a strong correlation of effect sizes (Pearson’s r = 0.76, *P* < 2.2 × 10^−16^), with 90.8% of associations maintaining consistent direction of effect. At FDR < 0.05, (*P* < 1.1 × 10^−5^), 99.3% (531,193) of our *cis*-meQTLs in EA replicated in the GoDMC EA cohort.

### Dissecting the drivers of population-specific methylation regulation

We observed substantial population-specific genetic regulation of DNAm. To ensure fair comparison across ancestries, we restricted a balanced sample size (N = 160) for both AA and EA cohorts in this analysis. First, we found that a significant fraction of this specificity was driven by ancestry-specific allele frequencies. Notably, 25.3% (12,817,961) of *cis*-meQTLs in the AA population were rare or absent in Europeans, only 5.7% (6,771,498) of EA *cis-*meQTLs were rare or absent in Africans. We defined rare or absent as two or fewer carriers in the corresponding ancestry in the Phase-3 1000 Genome Project^68^. This disparity was even more pronounced for sentinel *cis*-meQTLs, defined as the most significant SNP per CpG site, with 33.4% (494,316) of AA variants being rare or absent in Europeans compared to 11.1% (169,330) of EA variants in Africans (Fig. 2f).

We next investigate effect-size heterogeneity using Cochran’s Q test^82^. We reported *I*^2^ here, which quantifies the proportion of effect size variance attributable to true heterogeneity rather than sampling error. Among *cis*-meQTLs that were common in both populations (MAF > 5%), AA-specific *cis*-meQTLs (not in EA) exhibited high heterogeneity (mean *I*^2^ = 76.9%), with 36.1% reaching significant at FDR < 0.05 ((*Q*_*p*_ < 2.1 × 10^−4^), indicating genuine ancestry-specific regulatory effects. EA-specific associations showed lower overall heterogeneity (mean *I*^2^ = 53.4%), with 11.6% reaching (*Q*_*p*_ < 1.8 × 10^−4^ at FDR < 0.05 (Fig. 2g). Despite these differences, for shared variants, effect sizes showed high concordance between populations (*r* = 0.96,*P* < 2.2 × 10^−16^, Additional file 1: Fig. S9).

Together, these findings highlight that population-specific *cis*-meQTLs arise from a combination of allele frequency differences and true effect-size heterogeneity.

### *Cis*-h^2^ of CpG sites

To quantify genetic contributions to DNAm variation, we estimated *cis*-h^2^ (the proportion of methylation variation explained by *cis*-acting SNPs within 1 Mb) using REML in GCTA^63^. We observed that 52.0% and 58.2% of CpG sites exhibited *cis*-h^2^ < 0.01, 22.0% and 34.4% CpG sites with a *cis*-h^2^ between 0.01 and 0.1, and 26.0% and 7.4% CpG sites with a *cis*-h^2^ > 0.1 in the AA and EA populations, respectively (Fig. 3a). The average *cis*-h^2^ was significantly higher in the AA population (mean=0.08) compared to EA population (mean=0.03; *P*< 1 × 10^−16^ by Wilcoxon rank-sum test). We provide the estimated *cis*-h^2^ as valuable resources for the community to support subsequent analysis and CpG sites selection. Importantly, this ancestry difference in *cis*-h^2^ persisted when sample size was through down-sampling of EA (mean *cis*-h^2^ = 0.03), indicating that the higher *cis*-h^2^ in AA is not driven by sample size disparity, but reflects underlying genetic or regulatory differences between populations.

**Fig. 3.**
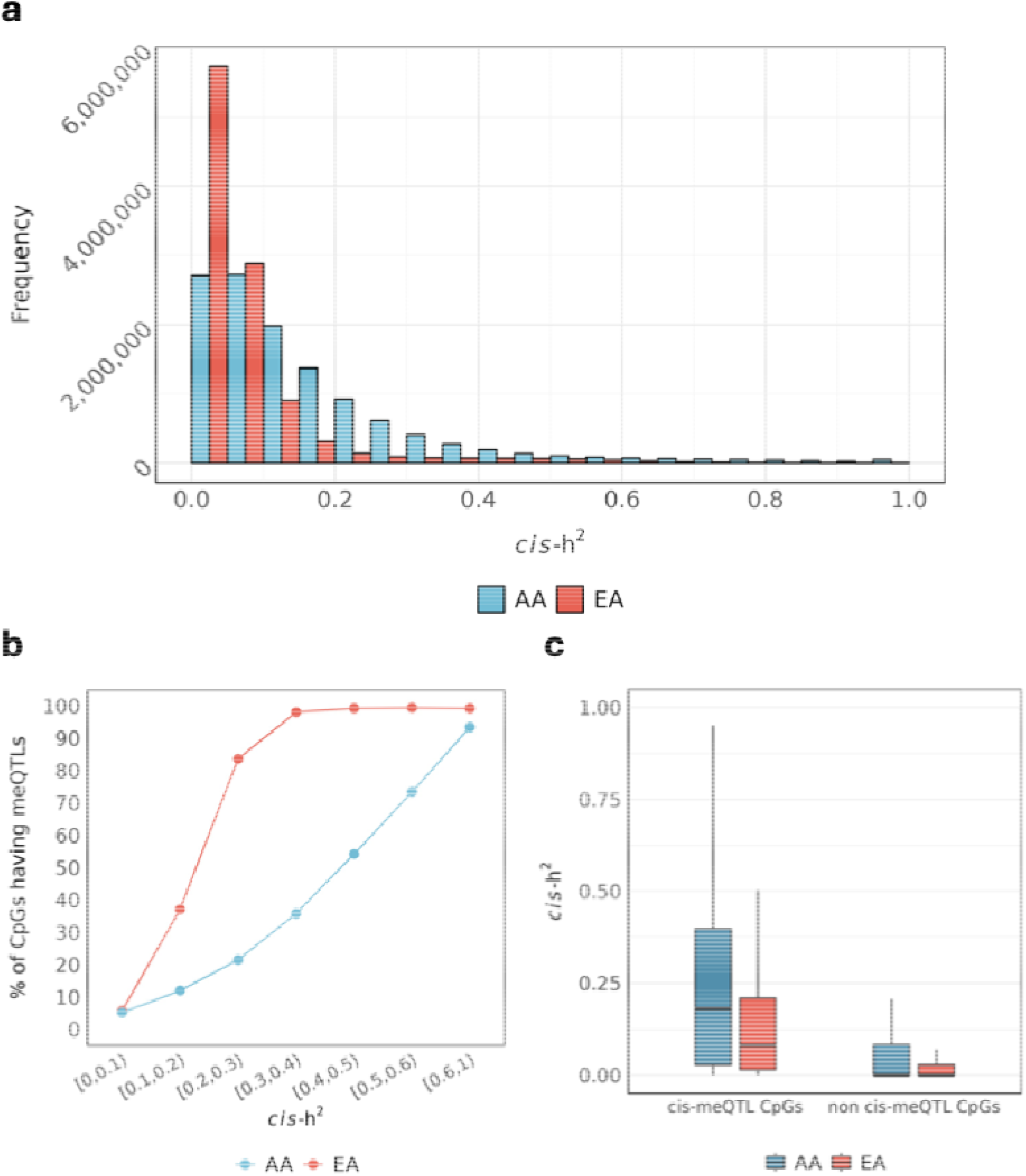
*cis*-h^2^. **(a)** Distribution of *cis*-heritability estimates across CpG sites with *cis*-h^2^ > 0.01 in AA and EA populations. **(b)** Relationship between CpG site *cis*-h^2^ and the presence of significant *cis*-meQTL associations, illustrating the concordance between heritability estimates and genetic regulation. **(c)** Comparative analysis of *cis*-h^2^ distribution among CpG sites with and without significant *cis*-meQTL in AA and EA ancestries.

As expected, integration of heritability estimates with *cis*-meQTL results revealed strong concordance. For highly heritable sites (*cis*-h^2^ > 0.5), the majority showed significant *cis*-meQTL associations (86.8% in AA, 99.4% in EA; Fig. 3b). CpG sites with identified *cis*-meQTLs exhibited substantially higher mean heritability (AA: 0.26, EA: 0.06) compared to those without (AA: 0.06, EA: 0.02; Fig. 3c).

We then compared *cis*-h2 estimates for CpGs captured by array-based 900K or 450K platforms with those derived from our full WGBS dataset. The distribution of *cis*-h2 values for CpG sites in the 450K and 900K sets closely mirrored that of the overall WGBS dataset (mean *cis*-h2 of 900K/450K: AA = 0.08/0.08, EA = 0.03/0.03; Additional file 1: Fig. S10), indicating that these platforms capture many highly regulated CpG sites. However, WGBS enables comprehensive genome-wide methylation profiling, allowing identification of additional heritable CpG sites and potentially novel regulatory regions not captured by targeted platforms.

### Multi-ancestry MWAS of complex traits in MVP

To illustrate the utility of our data source, we conducted MWAS. Specifically, we first built DNAm prediction models with *cis*-h^2^ > 0.01 for 2,677,714 CpG sites in AA population and 1,976,046 CpG sites in EA populations, with 1,075,702 CpG sites shared between both groups. The mean predictive performance (measured by R^2^) was 0.20 for AA and 0.18 for EA, outperforming models built on array-based CpG sites (450K/900K mean R^2^: AA=0.11/0.10; EA = 0.10/0.09). These findings align with the observed heritability patterns and demonstrating the enhanced predictive power enabled by WGBS-based comprehensive methylome coverage.

Using these prediction models, we performed MWAS for 41 complex traits using GWAS summary statistics from the MVP dataset^50^. After Bonferroni correction across all models and phenotypes (*P* < 4.7 × 10^−10^ for AA and *P* < 6.4 × 10^−10^ for EA), we identified 23,650 significant methylation-phenotype associations (Additional file 2: Tables S5.1 and S5.2), including 21,534 associations in EA and 2,116 in AA (Fig. 4a). The higher number of associations in EA likely reflects the larger sample size in EA GWAS data (mean = 261,202) compared to AA (mean = 54,271).

**Fig. 4.**
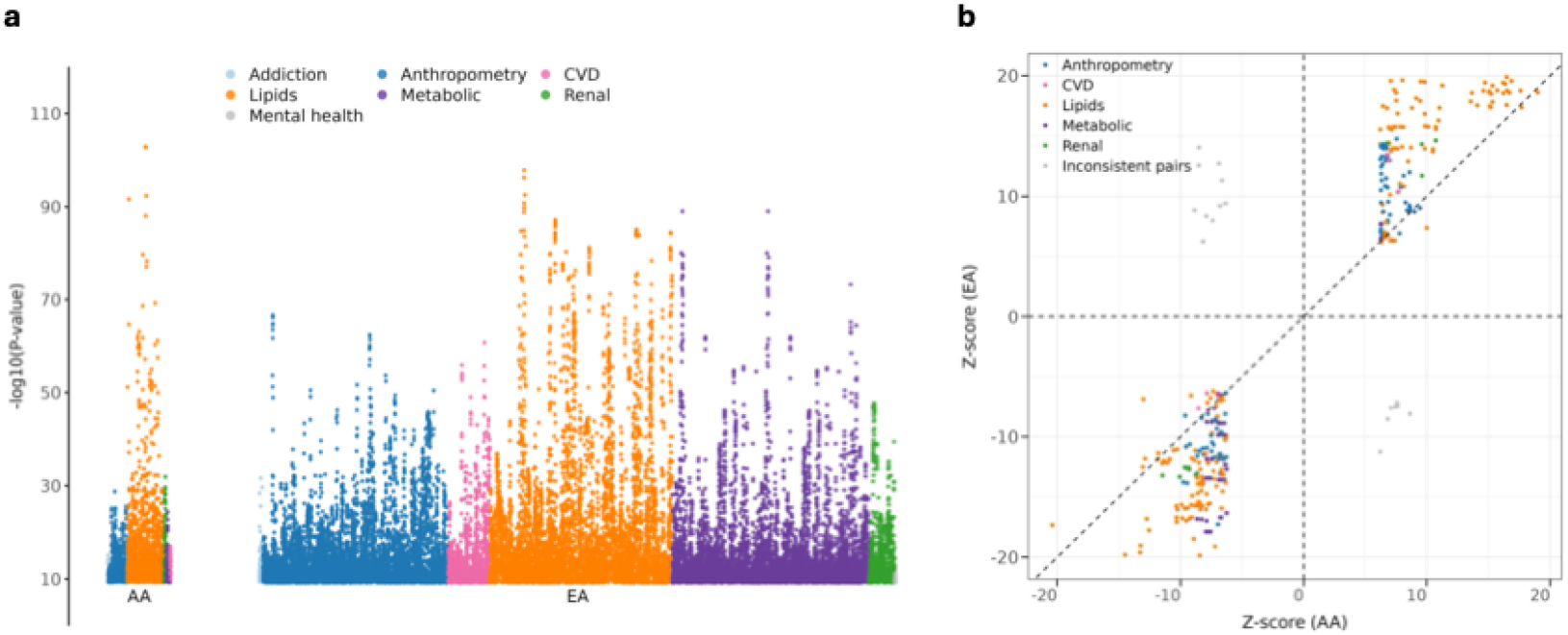
MWAS results for 41 phenotypes in the MVP dataset. **(a)** Distribution of significant MWAS associations after Bonferroni correction (*P* < 4.7 × 10^−10^ for AA and *P* < 6.4 × 10^−10^ for EA) across different trait categories. **(b)** Cross-ancestry comparison of effect sizes (Z-scores) for CpG-phenotype associations significant in both populations. Colored points represent consistent directional effects across ancestries, while grey points indicate discordant directions. Different colors denote distinct phenotype categories.

Notably, our WGBS-based approach uncovered associations not detectable with array-based platforms. For AA, 2,073 (98.0%) and 2,021 (97.5%) significant associations were not captured by the 450K and 900K methylation platforms, respectively. Similarly, for EA, 21,211 (98.5%) and 20,888 (97.0%) associations were beyond the coverage of these platforms. These results highlight the advantage of WGBS for capturing the full landscape of methylation-trait associations across ancestries.

Among the identified associations, 370 unique methylation-phenotype association pairs were significant in both ancestries; the majority (352 out of 370) of these associations displayed consistent directions of effects (Fig. 4b). This consistency suggests the presence of shared epigenetic mechanism across ancestries. These consistent associations encompassed 259 distinct CpG sites and spanned 10 phenotypes categorized into five broad groups: lipids, cardiovascular diseases (CVD), anthropometric measurements, metabolic conditions and renal traits. Genomic annotation revealed that most shared associations (321 of 352) involved CpG sites in open sea regions, with fewer sites in shores (16), shelves (10), and islands (5). This pattern highlights the regulatory potential of CpG sites in traditionally understudied genomic regions and reinforces the importance of comprehensive methylome coverage for discovering functional variations.

To aggregate CpG-level associations into gene-level insights, we mapped CpG sites from all associations to the genes and used Aggregated Cauchy Association Test (ACAT)^83^ to detect significant gene-phenotype associations.^84^ After Bonferroni correction (*P* < 2.5 × 10^−6^ for AA and *P* < 2.6 × 10^−6^ for EA), we identified 670 significant associations for AA population spanning 14 traits across 6 categories (Additional file 2: Table S6.1). We also identified 5,093 significant associations for EA population encompassing 27 traits and 7 categories (Additional file 2: Table S6.2). Among these, 468 associations are significant in both ancestries, covering 12 traits across 6 categories (Additional file 2: Table S7). Notably, the vast majority of gene-phenotype associations remained undetected by conventional methylation arrays: the 450K panel failed to capture 97.0% (650/670) of significant associations in the AA population and 97.3% (4,955/5,093) in the EA population. Similarly, the 900K panel missed 96.5% (647/670) and 96.6% (4,919/5,093) of significant associations in AA and EA populations, respectively. These findings highlight the substantial gain in discovery power offered by WGBS over array-based approaches.

### Novel T2D-associated methylation signatures and ancestry-specific regulatory mechanisms

Using our meQTL data and MWAS models, we conducted a comprehensive analysis of T2D. Our MWAS analysis identified 20 CpG sites significantly associated with T2D in the AA population and 4,023 CpG sites in the EA population after Bonferroni correction. Notably, 12 CpG sites showed consistent associations across both populations, suggesting shared epigenetic mechanisms underlying T2D pathophysiology.

To validate the causal nature of these associations, we performed MR analysis, which supported causal relationships for 75.0% (15/20) of significant CpG sites in AA and 72.0% (2,897/4,023) in EA after Bonferroni correction (*P* < 1.1 × 10^−9^ for AA and *P* < 2.2 × 10^−9^ for EA). Of the 12 cross-ancestry consistent associations, 83.3% (10/12) showed evidence of causality through MR. Complementary Bayesian colocalization analysis revealed that 35.0% (7/20) of AA and 29.6% (1,191/4,023) of EA T2D-associated CpG sites demonstrated strong evidence of shared genetic architecture with T2D GWAS signals (PPH4 > 0.8), indicating these epigenetic associations likely share causal variants with genetic associations.

When aggregating CpG-level associations to genes, we identified 13 genes significantly associated with T2D in both AA and EA populations (Additional file 2: Table S7). These findings point to immune-cell–specific epigenetic programs relevant to T2D pathogenesis.

Among cross-ancestry signals, *JAZF1*, a nuclear transcription regulator, has consistent T2D associations and acts as a mediator of metabolic stress by regulating ribosome biogenesis and global protein/insulin translation^85^. Critically, *JAZF1* also modulates T2D pathogenesis through monocyte-mediated mechanisms. In the immune compartment, obesity-driven inflammation is dominated by adipose-tissue macrophages (ATMs), where M1 polarization promotes insulin resistance^86^. *JAZF1* overexpression shifts macrophages toward an M2 phenotype, expands regulatory T cells, and lowers Tumor necrosis factor-alpha (TNF-α), Monocyte chemoattractant protein-1 (MCP-1), and IL-8^87^, with concordant reductions in CD11c^+^ macrophages and CD4^+^ T cells^88^. Consistent with these mechanisms, our purified-monocyte data reveal *JAZF1* methylation changes associated with T2D in both AA and EA cohorts, supporting epigenetic dysregulation of *JAZF1* in monocytes as a complement to its β-cell and adipose roles in T2D pathogenesis.

*GPSM1*, also shared across ancestries, aligns with a pro-inflammatory monocyte/macrophage program (e.g., Gαi/cAMP-PKA-CREB and TLR-NF-κB signaling) that is consistent with insulin-resistance biology^89^; its association here supports a model in which genetically driven methylation at *GPSM1* tunes inflammatory set-points in circulating monocytes. Canonical T2D loci (*TCF7L2, ANK1, FTO*) also appear in our gene-level aggregation; in monocytes, these signals plausibly converge on pathways influencing polarization, cytokine output, lipid handling, or trafficking, offering immune-cell explanations for genetic risk traditionally interpreted through β-cell or adipose biology^90–92^. We also identified novel associations with potential mechanistic roles: *NKX6-3*, which has not been directly associated with T2D risk in prior studies, is an islet β-cell transcription factor that is highly sensitive to oxidative stress. Because monocytes/macrophages are key responders to oxidative stress and can propagate pro-inflammatory signals that impair β-cell function, monocyte-localized methylation at *NKX6-3* may capture an immune–islet axis relevant to T2D pathogenesis.^93^

Notably, our analysis revealed six T2D-associated genes unique to the AA population. Among these, *BCKDHA*, encoding the branched-chain alpha-keto acid (BCAA) dehydrogenase (BCKD) subunit, is particularly interesting. This finding connects with previous observations of elevated BCAA levels in both human and animal models of obesity^94,95^, and the documented suppression of BCKD complex activity in liver and adipose tissue during obesity^96^. Given BCKD’s expression across multiple tissues including monocytes, this ancestry-specific association suggests potential immune system involvement in T2D pathogenesis that may be particularly relevant in AA populations.

Comparison with array-based approaches demonstrated the superior detection power of WGBS at both CpG site and gene levels. At the CpG site level, conventional methylation arrays captured only 5.0% (1/20) and 2.4% (97/4,023) of the significant T2D-associated sites identified by WGBS in AA and EA populations, respectively. At the gene level, the limitation of array-based approaches was particularly evident in T2D analysis, where arrays identified only three genes (*TCF7L2, FTO*, and *KCNC2*) in both ancestries, missing numerous biologically relevant associations detected through WGBS, as discussed above.

### Identification of *trans*-meQTL for whole genome DNAm sites across two populations

Given our modest sample sizes (AA: n=160; EA: n=298), we conducted an exploratory analysis of *trans*-meQTL associations to gain preliminary insights into long-range methylation regulation. Controlling FDR at 0.01, our analysis revealed 11,793,417 *trans*-meQTL associations (*P*< 3.7 × 10^−10^) involving 608,243 CpG sites (2.4% of all tested sites) in the AA population, and 51,883,518 *trans*-meQTL associations (*P* < 4.7 × 10^−11^) involving 1,070,366 CpG sites (4.3% of all tested sites) in the EA population. Notably, we observed minimal overlap between populations, with only 4,368 shared CpG sites showing *trans*-meQTL associations. The majority of associations were population-specific, with 97.8% unique to AA and 96.9% unique to EA populations. Under a more liberal threshold of *P*< 1 × 10^−s^, our analysis yielded 38,007,510 *trans*-meQTLs in AA (involving 1,727,614 CpG sites) and 79,796,981 *trans*-meQTLs (involving 1,834,716 CpG sites) in EA, with 5,989 shared CpG sites in both ancestries. Note that these patterns should be interpreted cautiously given the sample size limitations.

Initial characterization of these putative *trans*-meQTL associations revealed several patterns that warrant further investigation in larger cohorts. We observed a negative correlation between

MAF and effect size for variants with MAF > 0.05 (AA: correlation = -0.18, *P* = 2.2 × 10^−16^, EA: correlation = -0.38, *P* < 2.2 × 10^−16^), consistent with power/selection effects at lower MAF (Fig. 5a). The relationship between effect size and genomic distance showed stronger distance-dependent attenuation for *trans*-meQTLs compared to *cis*-meQTLs (Fig. 5b). We observed high concordance of effect sizes of shared *trans*-meQTLs between populations (r = 0.97, *P*< 2.2 × 10^−16^, Fig. 5c). When considering all tested pairs, correlation was modest (r = 0.64, *P*< 2.2 × 10^−16^); among associations significant in at least one ancestry, correlation improved substantially improved (r = 0.87, *P* < 2.2 × 10^−16^; Additional file 1: Fig. S11). This indicates that the most robust *trans*-meQTLs exhibit high cross-population consistency.

**Fig. 5.**
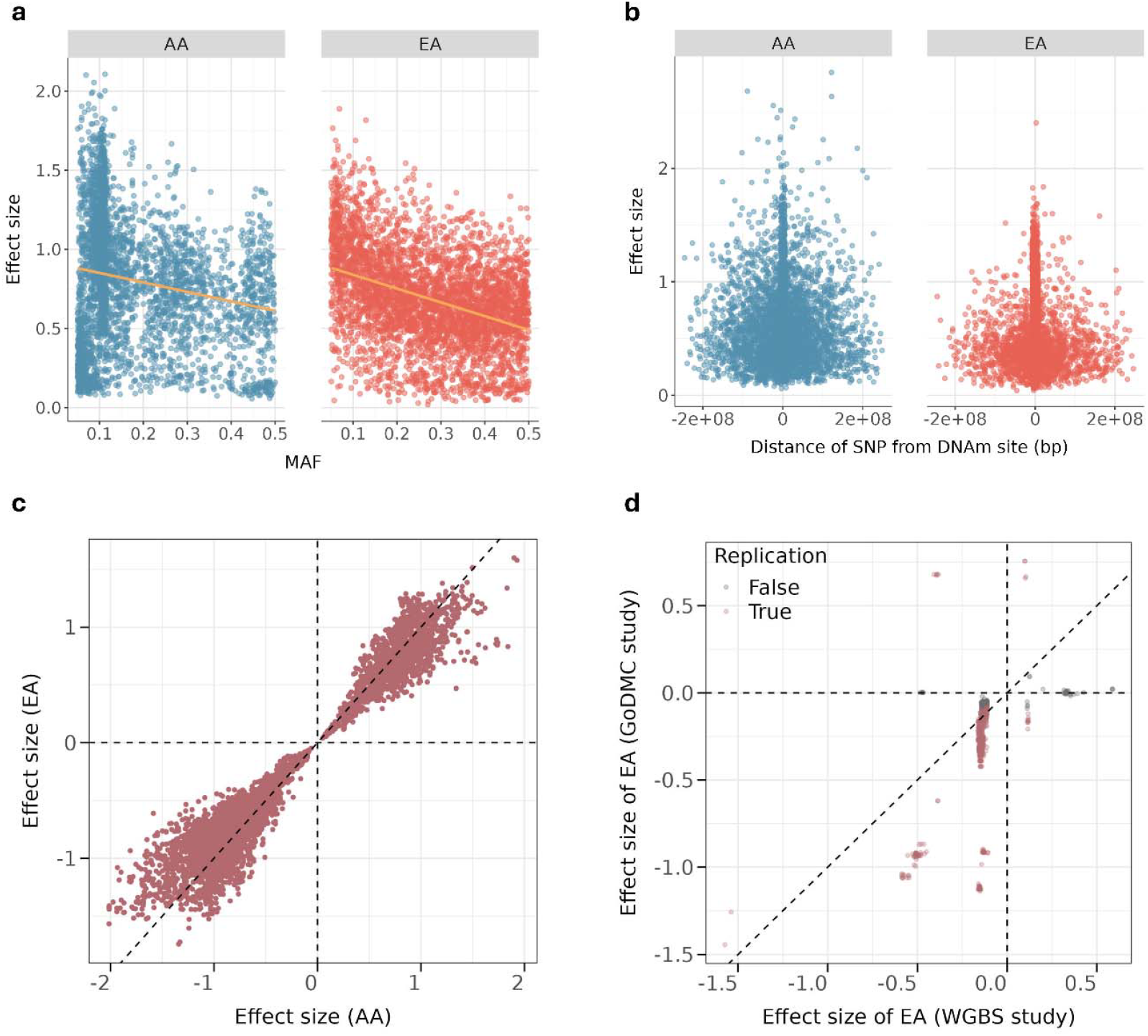
Characterization of *trans*-meQTL effects across African American (AA) and European American (EA) populations. **(a)** Relationship between *trans*-meQTL effect sizes and MAF, demonstrating the impact of allele frequency on long-range methylation. Orange regression lines indicate overall trends. **(b)** Effect sizes distribution as a function of genomic distance between SNP and DNAm site in AA (left) and EA (right). **(c)** Correlation of *trans*-meQTL effect sizes between AA and EA populations for shared associations, demonstrating the consistency of *trans*-effect across ancestries. **(d)** Replication effect sizes of WGBS study (EA, n = 298) and GoDMC study (EA, n = 27,750).

To validate our *trans*-meQTLs, we performed a replication analysis using the GoDMC dataset in EA. When restricting to 46,114 CpG sites that are available in both studies, our study identified 906,738 significant *trans*-meQTL. Among these, only 1,033 (0.1%) were also present among the 8,713,505 *trans*-pairs reported in the GoDMC cohort, largely reflecting the limited overlap between WGBS and the 450K array used in GoDMC (which captures only ∼1.5% of CpG sites measured by WGBS) and the fact that data shared by GoDMC had been pre-filtered, retaining only meQTLs that met a *P*< 1 × 10^−5^ threshold in an individual study from the meta-analysis. Despite this small overlap, we found that 82.2% (849 of 1,033) of these shared associations were also significant in the GoDMC cohort (FDR < 0.05, *P* < 3.5 × 10^−10^). Furthermore, these associations showed high directional consistency (96.5%) and a strong positive correlation in effect sizes (Pearson’s r = 0.51; Fig. 5d). This pattern of high concordance within a limited overlap confirms the replicability of our *trans*-meQTL findings.

## Discussion

We present the first comprehensive analysis of genetic influences on monocyte DNAm across AA and EA populations using WGBS. This study contributes several key advances to the field of epigenetics. First, by analyzing over 25 million CpG sites, we identified approximately ten times more *cis*-meQTL associations than previous studies^11^, providing unprecedented resolution into the regulation of DNAm. Second, our population-specific DNAm imputation models, validated using the MVP dataset across 41 complex traits, demonstrate the potential of integrative MWAS to complement GWAS in identifying trait-associated CpG sites and genes that may offer insights into disease biology and potential therapeutic avenues. Third, our use of purified monocytes enables for precise characterization of cell-type-specific methylation patterns, which is critical for understanding immune-related diseases such as T2D.

Our cross-ancestry analysis offers valuable insights into population-specific methylation regulation. Notably, the AA population showed higher *cis*-h^2^ of DNAm levels compared to EA. Furthermore, approximately 25.3% of *cis-*meQTLs identified in the AA population were rare or absent in Europeans, whereas only 5.7% of EA-specific *cis-*meQTLs were rare in Africans. We also observed substantial true effect-size heterogeneity. Among shared common variants, AA-specific *cis*-meQTLs showed markedly higher heterogeneity (mean *I*^2^ = 76.9%; 36.1% significant at FDR < 0.05) than EA-specific associations (mean *I*^2^ = 53.4%; 11.6% significant), indicating genuine ancestry-driven regulatory differences beyond allele frequency. These findings demonstrate that ancestry-specific meQTLs arise from both genetic architecture and context-dependent regulatory effects.

The comprehensive coverage (over 25 million CpG sites) of WGBS enabled us to uncover numerous methylation markers beyond the scope of conventional array-based approaches. Our replication analysis against large whole blood cohorts (GoDMC, GENOA) highlights the value of this approach. While the high concordance of effect sizes for shared CpGs validates our findings and indicates that many meQTLs are shared across blood cell types, this comparison is limited to the ∼1.5-3% of CpG sites captured by array platforms. This limitation underscores why whole blood studies are not sufficient. First, they leave the genetic regulation of the vast majority of the methylome uncharacterized. Second, whole blood is a heterogeneous mixture, and associations cannot be mechanistically resolved to a specific cell type. For example, our analysis not only replicated general T2D-associated genes such as *JAZF1* and *GPSM1*, but also provides a specific mechanistic link, showing that their methylation changes in monocytes are consistent with inflammatory mechanisms of insulin resistance. Such cell-type-specific insights are obscured in whole blood analyses. Furthermore, our WGBS approach also identified novel, ancestry-specific associations (e.g., *BCKDHA* in the AA population). This limitation underscores why current whole blood-based studies are not sufficient.

Our findings align with recent studies emphasizing that the DNAm landscape is strongly influenced by cell-type-specific regulatory programs^97^. While cell-type identity establishes the baseline methylation framework, we demonstrate that genetic variation plays a crucial role in fine-tuning these patterns, even within a homogeneous cell population like monocytes. This suggests a hierarchical model of methylation regulation where cell-type programs establish the foundational pattern, which is then modulated by genetic factors in a population-specific manner. Our analysis provides direct evidence for this model, revealing how genetic variation can influence methylation patterns within a defined cellular context.

Several limitations of our study merit discussion and offer directions for future work. First, the relatively modest sample sizes, particularly in the AA cohort, may have led to an underestimation of *cis*-meQTL associations and limited power to detect *trans*-effects, which typically have smaller effect sizes and require larger samples to identify^98^. This limitation also explains the limited overlap observed when comparing our *trans*-meQTL results with the GoDMC whole blood study (with only 1,033 shared). Nonetheless, this remains one of the largest WGBS studies to date, particularly in its inclusion of both EA and AA populations. Second, the male-predominant composition of our cohort (all males), as well as the MVP (∼90% male) dataset may obscure sex-specific methylation differences. Although DNAm patterns are largely consistent across sexes^99^, subtle sex-specific effects may still exist.^100^ Third, while our focus on purified monocytes provides valuable insights into immune cell regulation, future studies should expand to other cell types and integrate transcriptomic data to build a more holistic view of epigenetic regulation. Additionally, donor cytomegalovirus (CMV) serostatus was not assessed; given the potential impact of latent CMV on circulating immune phenotypes, including monocytes, residual confounding cannot be excluded and should be addressed in future cohorts via CMV serology and/or viral load measurement. Overall, our work lays the foundation for broader exploration of tissue- and context-specific methylation landscapes and their roles in complex traits and diseases.

## Conclusions

We build a new, comprehensive resource of monocyte methylome across European and African populations. Based on this resource, we performed meQTL analyses, investigated the reasons underlying ancestry-specific results, built DNAm imputation models, and conducted MWAS of 41 traits in MVP. These resources provided new insights into the etiology of type 2 diabetes and we expect them to be used in other diseases.

## Data Availability

All data produced are available online at www.gcbhub.org and https://osf.io/gct57/
All codes are available online at https://github.com/ChongWuLab/Multi-ancestry-monocytes-methylome-study

https://www.gcbhub.org/phenotypes_MWAS

https://osf.io/gct57/

## List of abbreviations

AA: African Americans
ACAT: Aggregated Cauchy Association Test
ATMs: Adipose-Tissue Macrophages
BCAA: Branched-Chain Alpha-Keto Acid
BCKD: Branched-Chain Alpha-Keto Acid Dehydrogenase
BMI: Body Mass Index
BSA: Bovine Serum Albumin
BWA: Burrows-Wheeler Aligner
*cis*-h2: *cis*-heritability
CMV: Cytomegalovirus
CVD: Cardiovascular Diseases
DNAm: DNA Methylation
DNBs: DNA Nanoballs
EA: European Americans
ECDF: Empirical Cumulative Distribution Function
FDR: False Discovery Rate
GATK: Genome Analysis Toolkit
GCTA: Genome-wide Complex Trait Analysis
GENOA: Genetic Epidemiology Network of Arteriopathy
GoDMC: Genetics of DNA Methylation Consortium
GRM: Genetic Relationship Matrix
GWAS: Genome-Wide Association Studies
HIV: Human Immunodeficiency Virus
Indel: Insertion/Deletion
INT: Inverse Normal Transformation
IQR: Interquartile Range
LD: Linkage Disequilibrium
LM-PCR: Ligation-Mediated Polymerase Chain Reaction
LOS: Louisiana Osteoporosis Study
MAF: Minor Allele Frequency
MCP-1: Monocyte Chemoattractant Protein-1
meQTL: Methylation Quantitative Trait Loci
MR: Mendelian Randomization
MVP: Million Veteran Program
MWAS: Methylome-Wide Association Studies
PBMCs: Peripheral Blood Mononuclear Cells
PC: Principal Components
PCA: Principal Component Analysis
PPH4: Posterior Probability of a shared causal variant
QC: Quality Control
REML: Restricted Maximum Likelihood
scRNA-seq: Single-Cell RNA-Sequencing
SNP: Single Nucleotide Polymorphism
T2D: Type 2 Diabetes
TNF-α: Tumor Necrosis Factor-alpha
VQSR: Variant Quality Score Recalibration
WGBS: Whole-Genome Bisulfite Sequencing
WGS: Whole-Genome Sequencing

## Declarations

### Ethics approval and consent to participate

The study was approved by the Tulane University Institutional Review Board (IRB #: 10-184088) and conducted in accordance with the principles of the Declaration of Helsinki. All recruited individuals signed an informed consent document prior to data and biosample collection.

### Availability of data and materials

All meQTL results, MWAS models, and associated analysis resources are publicly available at https://osf.io/gct57/. All codes are publicly available at https://github.com/ChongWuLab/Multi-ancestry-monocytes-methylome-study.

### Consent for publication

Not applicable.

### Authors’ contributions

H.W.D., C.W., and H.S. designed the study. C.Q., X.Z., K.J.S., Z.L., and Q.T. collected and preprocessed the whole genome bisulfite sequencing and whole genome sequencing data. W.Z. conducted the main analysis and wrote the first draft of the manuscript. Z.Z. created the online interactive website for sharing the results. M.L., B.Z., and L.W. provided insightful suggestions and contributed to the study design. All authors read, revised, and approved the final manuscript.

## Acknowledgements

We thank all participants of the Louisiana Osteoporosis Study (LOS) for their invaluable contributions. We are also grateful to the editor and the two anonymous reviewers for their constructive comments, which substantially improved this work.

## Additional files

Additional file 1: Supplementary figures

Additional file 2: Supplementary tables

