## Supplemental Figures for "An atlas of genetic effects on the monocyte methylome across European and African populations"

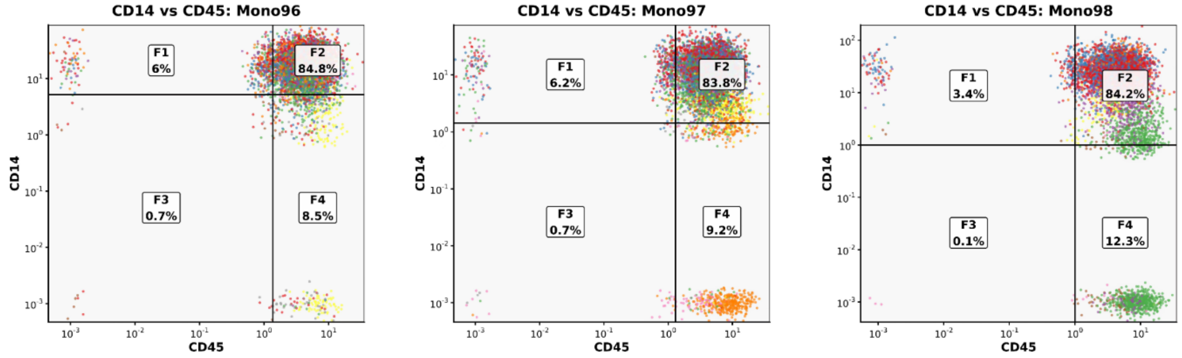


**Fig. S1: Proportion of CD14$^+$/CD45$^+$ cells identified by scRNA-seq–based cell-type clustering in three enriched human PBM samples.** CD14 and CD45 are the specific membrane markers on monocytes and mononuclear cells, respectively.

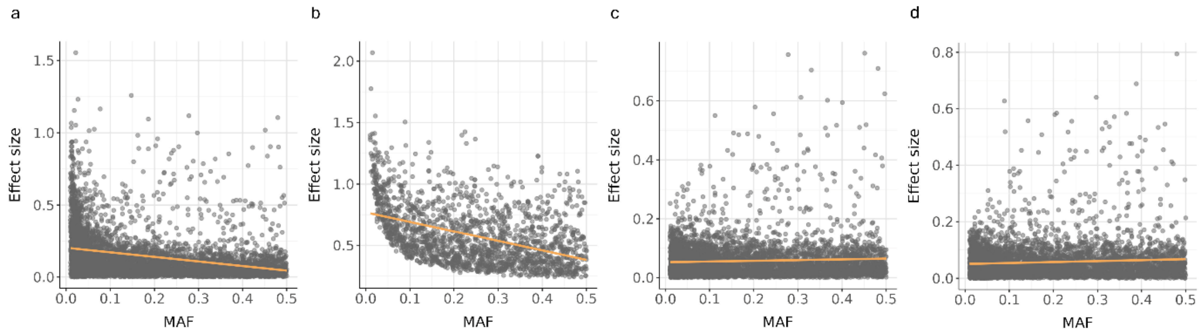


**Fig. S2:** Simulation shows the selection-driven MAF-effect-size pattern (a) All SNP-CpG tests; (b) Significant pairs (FDR < 0.05) (c) All tests with standardized effects. (d) Significant pairs with standardized effects (FDR < 0.05).

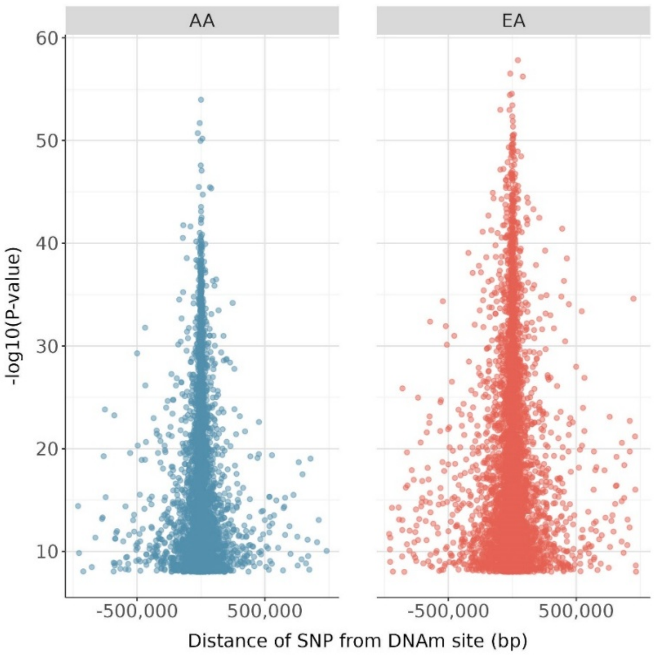


**Fig. S3: Relationship between p-values and the distance from SNP to DNAm site in the AA and EA populations.** The scatter plots depict the association between the distance from the SNP to the DNAm site (measured in base pairs, bp) and the corresponding -log10(p-values) for the detected meQTLs in the AA (left panel) and EA (right panel) populations. Higher values indicate more significant meQTL associations.

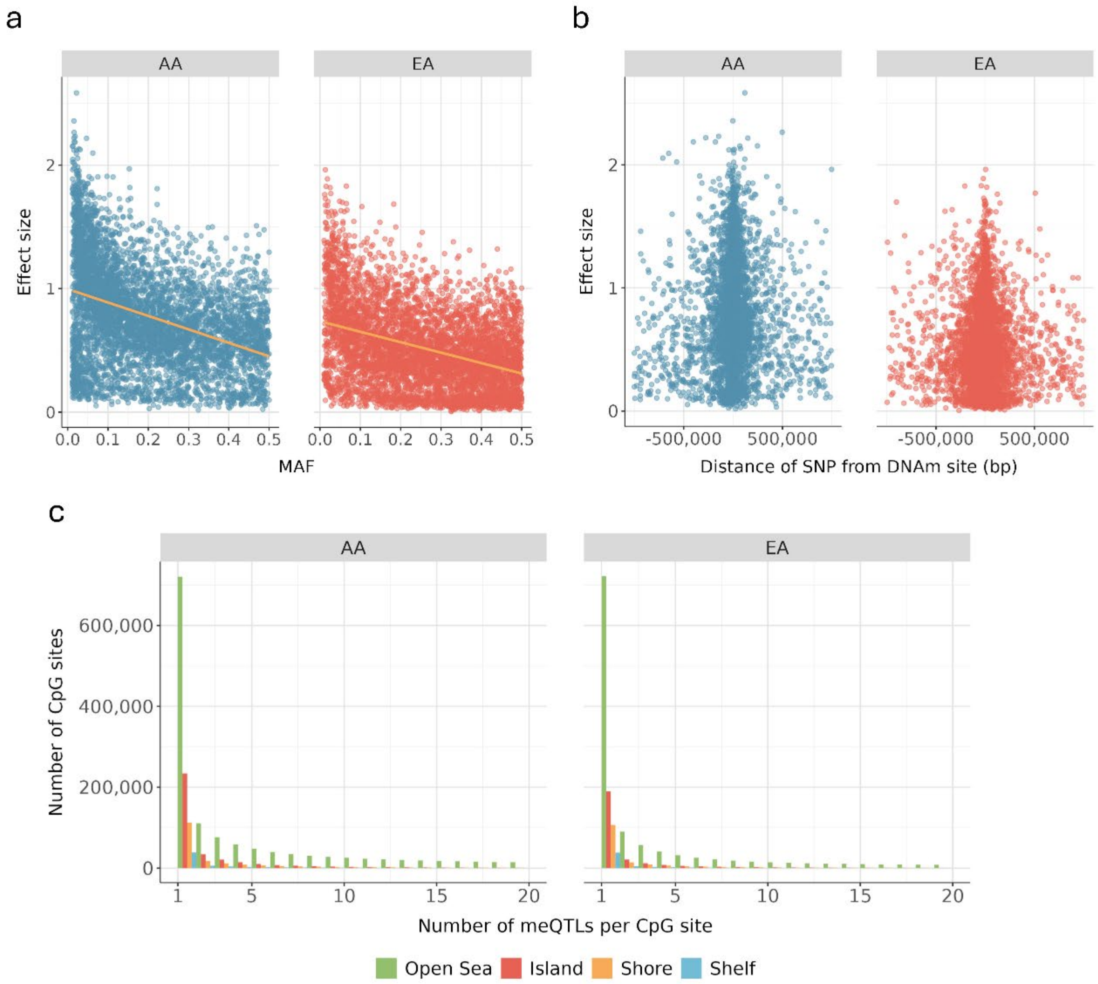


**Fig. S4: Characterization of *cis*-meQTLs using a MAF ≥ 1% cutoff. (a)** Relationship between *cis*-meQTL effect sizes and MAF with regression trends shown in orange. **(b)** Effect size distribution as a function of distance between SNP and DNAm site in AA (left) and EA (right) populations, measured in base pairs. **(c)** Genomic distribution of CpG sites with cis-meQTLs. The bar chart categorizes CpG sites based on the number of cis-meQTLs identified, with annotations for four genomic regions: CpG islands, which are regions with a high CpG density; CpG shores, located within 2 kb of islands; CpG shelves, extending an additional 2 kb from shores; and Open Sea, which represents areas more distal to the islands, shores, and shelves. Each region is color-coded.

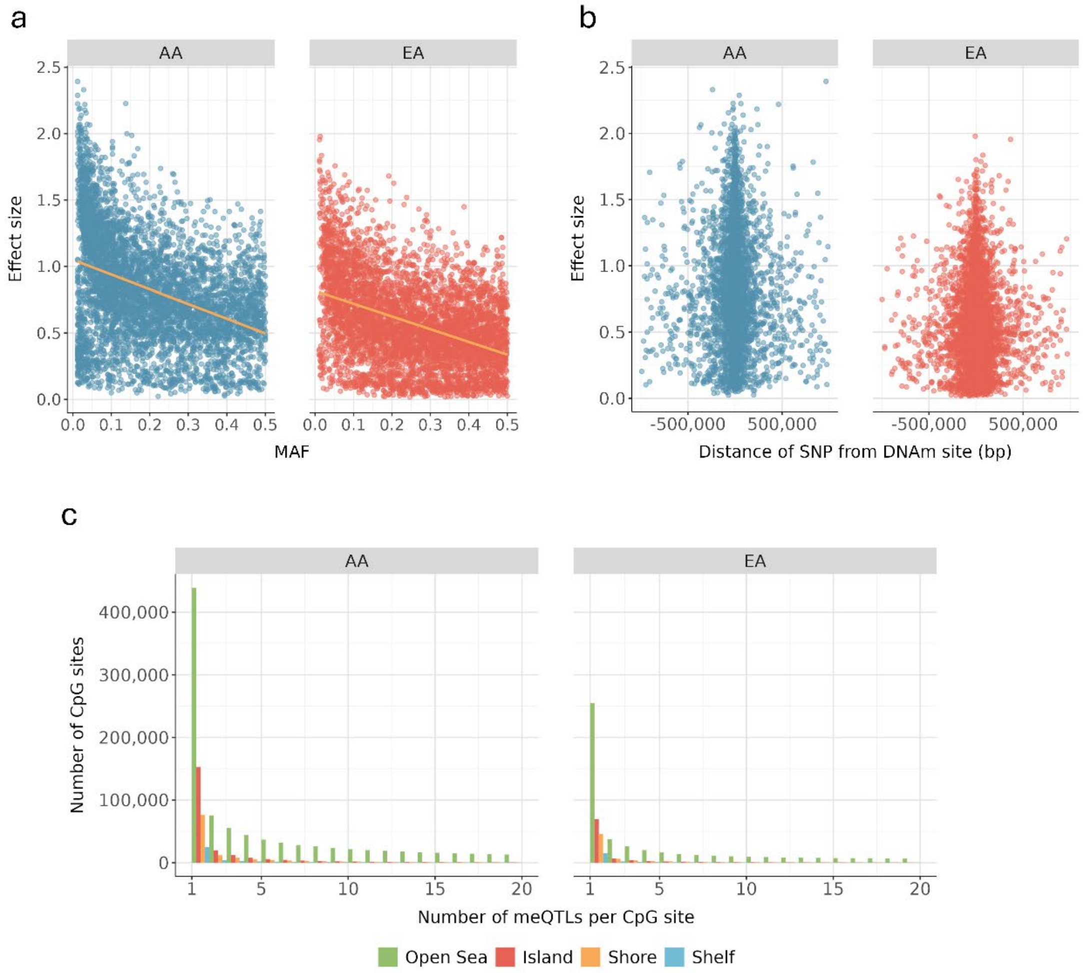


**Fig. S5: Characterization of *cis*-meQTLs using a fixed $P < 1 \times 10^{-6}$ cutoff. (a)** Relationship between *cis*-meQTL effect sizes and MAF with regression trends shown in orange. **(b)** Effect size distribution as a function of distance between SNP and DNAm site in AA (left) and EA (right) populations, measured in base pairs. **(c)** Genomic distribution of CpG sites with cis-meQTLs. The bar chart categorizes CpG sites based on the number of cis-meQTLs identified, with annotations for four genomic regions: CpG islands, which are regions with a high CpG density; CpG shores, located within 2 kb of islands; CpG shelves, extending an additional 2 kb from shores; and Open Sea, which represents areas more distal to the islands, shores, and shelves. Each region is color-coded.

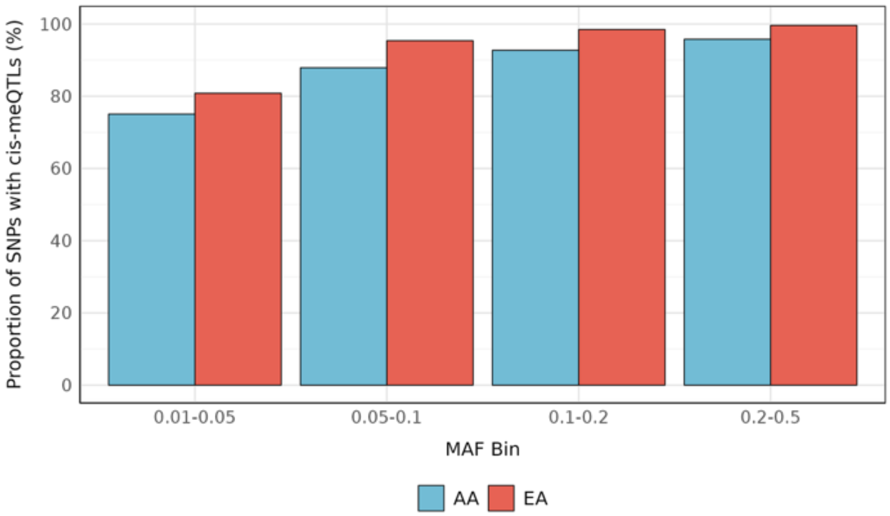


**Fig. S6: Proportion of variants with cis-meQTLs ($P < 1 \times 10^{-6}$) per MAF bin.**

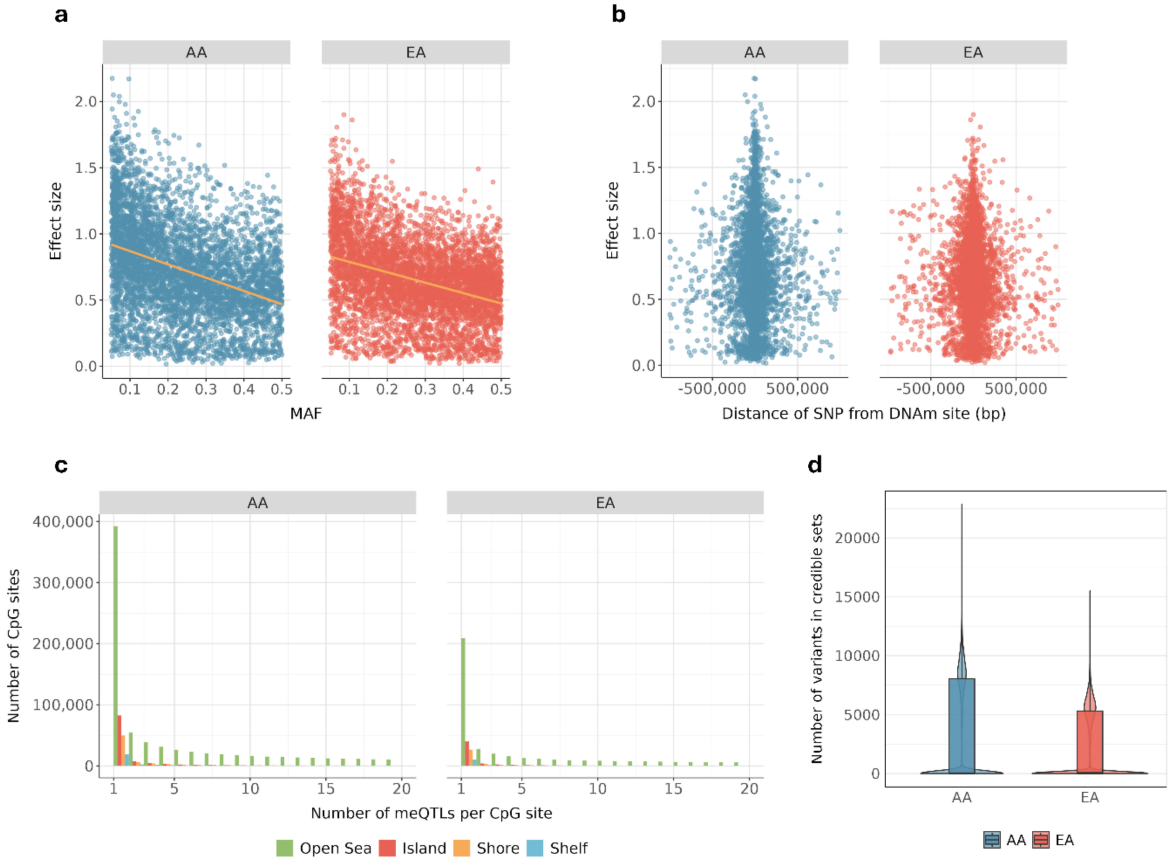


**Fig. S7: Characterization of *cis*-meQTLs for down-sampled EA (n = 160). (a)** Relationship between *cis*-meQTL effect sizes and minor allele frequencies MAF with regression trends shown in orange. **(b)** Effect size distribution as a function of distance between SNP and DNAm site in AA (left) and EA (right) populations, measured in base pairs. **(c)** Genomic distribution of CpG sites with cis-meQTLs. The bar chart categorizes CpG sites based on the number of cis-meQTLs identified, with annotations for four genomic regions: CpG islands, which are regions with a high CpG density; CpG shores, located within 2 kb of islands; CpG shelves, extending an additional 2 kb from shores; and Open Sea, which represents areas more distal to the islands, shores, and shelves. Each region is color-coded. **(d)** Fine-mapping results showing the distribution of credible set sizes from SuSiE analysis for CpG sites with significant *cis*-meQTLs in both populations. Box plots show median, quartiles, and 1.5×IQR whiskers; violin width indicates data density.

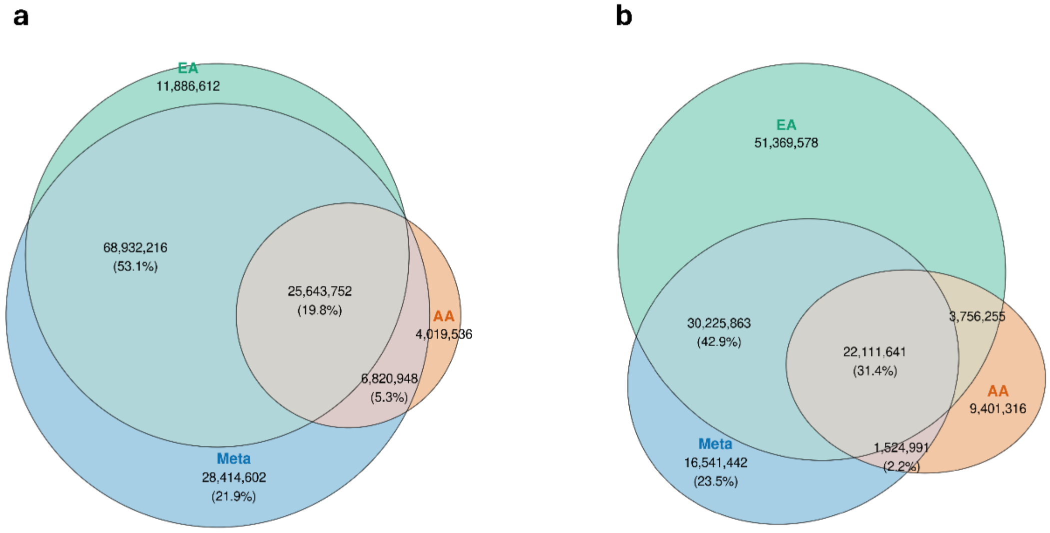


**Fig. S8: Venn diagrams showing overlap of significant *cis*-meQTLs (FDR < 0.01) between AA, EA, and meta-analysis. (a)** Fixed-effects meta-analysis. **(b)** Random-effects meta-analysis. All numbers indicate the count of significant associations in each region. Percentages shown within the meta-analysis (blue) regions indicate their proportion relative to the total number of meta-analysis associations.

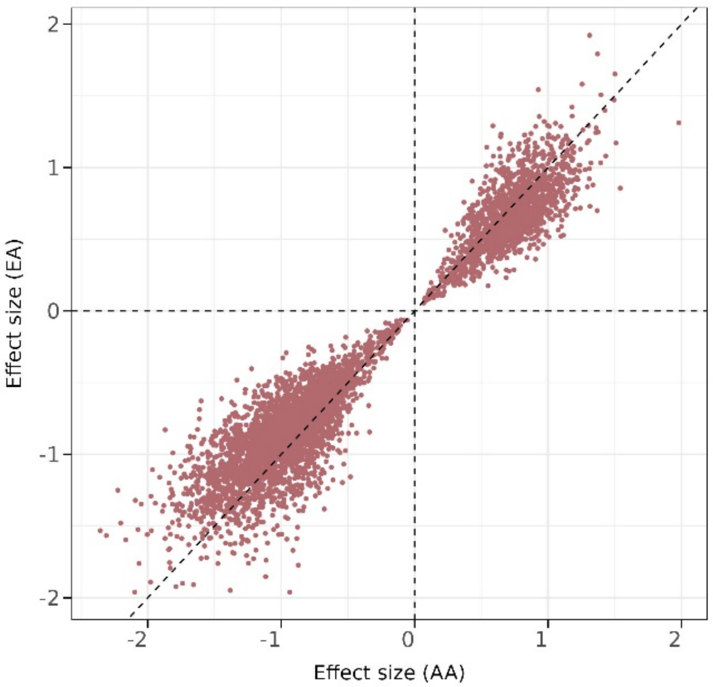


**Fig. S9 Correlation of cis-meQTL effect sizes between AA and EA population.**

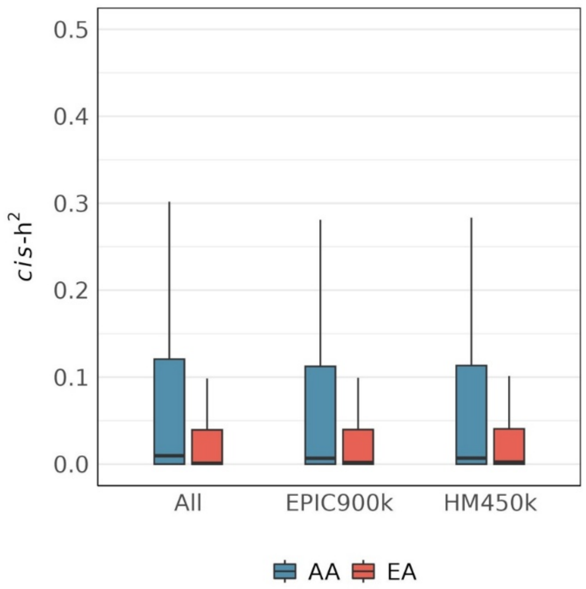


**Fig. S10** Comparison of *cis*-h2 distributions between WGBS data and conventional array-based platforms (900K and 450K), highlighting the comprehensive coverage of WGBS. Box plots show median, quartiles, and 1.5×IQR whiskers.

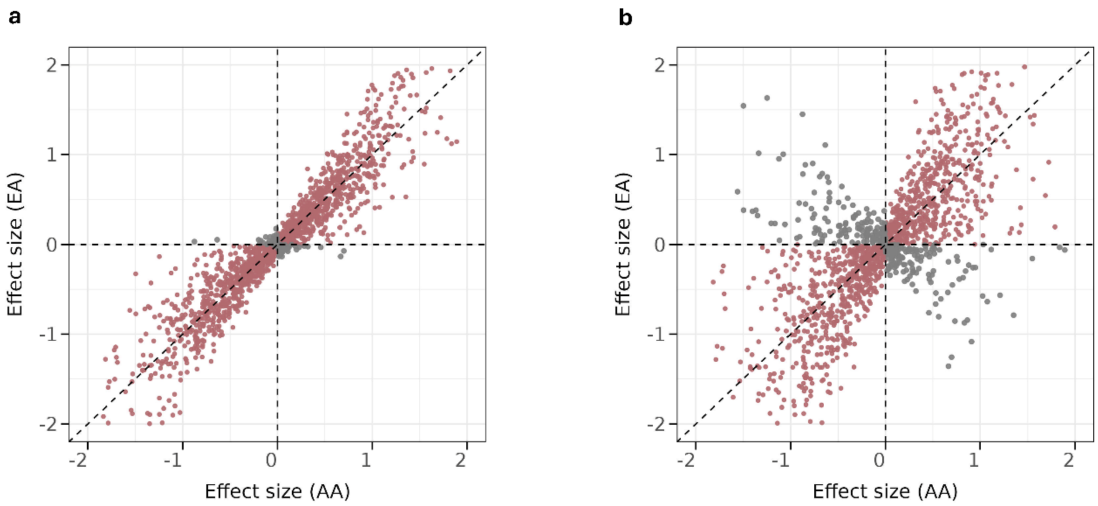


**Fig. S11: Correlation of *trans*-meQTL effect sizes between AA and EA population. (a)** Significant *trans*-meQTL in either population. **(b)** All tested CpG-SNP pairs.

### Note S1: Evaluation and filtering of CpG sites overlapping SNP positions

SNPs within CpG dinucleotides (CpG–SNPs) can distort bisulfite-based DNAm measurements or induce non-biological genotype–methylation associations.

Across the profiled methylome, we identified 3.8% of CpG sites (970,539 out of 25,721,231) in AA and 2.4% of CpG sites (607,275 out of 25,721,231) in EA that overlap with known SNP positions (MAF > 0.01). These CpG–SNP sites exhibited substantially inflated meQTL characteristics compared with non-overlapping sites:

- In AA, 82.1% (796,367 out of 970,539) of the CpG-SNP sites had at least one *cis*-meQTL, compared to only 10.4% (2,569,292 out of 24,750,692) of the non-CpG-SNP sites. The pattern is similar in EA population, with 91.0% (552,456 out of 607,275) of the CpG-SNP sites had at least one *cis*-meQTL, compared to only 9.5% of the non-CpG-SNP sites.
- In AA, these CpG-SNP sites were associated with an average of 55.7 SNPs, while the non-CpG-SNP sites were associated with only 24.2 SNPs on average. In EA, CpG-SNP sites were associated with an average of 151.0 SNPs, while the non-CpG-SNP sites were associated with only 54.6 SNPs.
- The mean estimated *cis*-$h^2$ of CpG-SNP sites is 0.40 in AA and 0.31 in EA, which is higher than the mean estimated cis-heritability of non-CpG-SNP sites (0.08 in AA and 0.03 in EA).

Together, these patterns are most parsimoniously explained by allelic mapping/measurement artifacts at the CpG–SNP locus rather than true biological effects.
